# COVID-19 vaccination atlas: an integrative systems vaccinology approach

**DOI:** 10.1101/2024.05.22.24307755

**Authors:** Wasim Aluísio Prates-Syed, Dennyson Leandro Mathias da Fonseca, Shahab Zaki Pour, Lena F Schimke, Aline Lira, Nelson Cortes, Jaqueline Dinis Queiroz Silva, Evelyn Carvalho, Igor Salerno Filgueiras, Tania Geraldine Churascari Vinces, Lorena C. S. Chaves, Gerhard Wunderlich, Ricardo Durães-Carvalho, Niels O. S. Câmara, Haroldo Dutra Dias, Hans D. Ochs, José E. Krieger, Helder I Nakaya, Otávio Cabral-Marques, Gustavo Cabral-Miranda

**Affiliations:** Department of Immunology, Institute of Biomedical Sciences (ICB), University of São Paulo (USP), São Paulo, Brazil; The Interunits Graduate Program in Biotechnology of the University of São Paulo, the Butantan Institute, and the Technological Research Institute of the State of São Paulo, Brazil; Interunit Postgraduate Program on Bioinformatics, Institute of Mathematics and Statistics (IME), University of Sao Paulo (USP), Sao Paulo, Brazil; Laboratory of Molecular Evolution and Bioinformatics, Department of Microbiology, Biomedical Sciences Institute, University of São Paulo, São Paulo, Brazil; Department of Parasitology, Institute of Biomedical Sciences (ICB), University of São Paulo (USP), São Paulo, Brazil; Department of Clinical and Toxicological Analyses, School of Pharmaceutical Sciences, University of São Paulo, São Paulo, Brazil; Department of Microbiology and Immunology, School of Medicine, Emory University, Atlanta, Georgia, United States; São Paulo School of Medicine, Department of Microbiology, Immunology and Parasitology, and Department of Morphology and Genetics, Federal University of São Paulo (UNIFESP), São Paulo, SP, Brazil; Department of Infectious and Parasitic Diseases, Faculty of Medicine, University of São Paulo, São Paulo, Brazil; Heart Institute, Clinical Hospital, Faculty of Medicine, University of São Paulo, Brazil; Laboratory of Genetics and Molecular Cardiology, Clinical Hospital, Faculty of Medicine, University of São Paulo, Brazil; Department of Pediatrics, University of Washington School of Medicine, and Seattle Children’s Research Institute, Seattle, WA, USA; Department of Neuroscience, Institute of Biomedical Sciences, Federal University of Minas Gerais (UFMG), Belo Horizonte, Brazil; Hospital Israelita Albert Einstein, São Paulo, SP, Brazil; DO’R Institute for research, São Paulo, Brazil; Department of Medicine, Division of Molecular Medicine, University of São Paulo School of Medicine, São Paulo, Brazil; Institute of Tropical Medicine, Faculty of Medicine of the University of São Paulo, Brazil

## Abstract

The COVID-19 vaccinations have played a significant role in controlling the pandemic. To elucidate their impact on the immune system, a COVID-19 vaccination atlas was developed through an integrative systems vaccinology approach. The atlas includes both healthy individuals and those infected with or without prior vaccination, and covers the administration of five vaccines in different regimens: Covilo®, Zifivax®, Vaxzebria® or Covishield®, Spikevax®, and Comirnaty®. Critical markers were identified to discriminate the different types of vaccines and infection, in which infection was associated with GATA3, ZNF3, KMT2A, ASXL1, SP100, and GZMM, and vaccine types were marked by ITGAM, ACTG1, LGALS3, and STAT5B. Additionally, the immunological signatures of heterologous vaccination and infection were described, and it was also shown how a full vaccination regimen markedly limited the shift of immune responses during natural infection, thereby constraining disease progression. Finally, the common transcripts shared across COVID-19 vaccines and vaccines against other pathogens were described.

## 1. Introduction

Coronavirus Disease 2019 (COVID-19) is a respiratory disease caused by Severe Acute Respiratory Syndrome Coronavirus 2 (SARS-CoV-2), with its first documented outbreak in Wuhan, China, in 2019 ^1^. The emergence of variants of concern (VOCs) urged adjustments in strategies to effectively control their spread due to the heterogeneity of symptoms, severity, transmission, and decreased efficacy of vaccines against new VOCs, especially the Omicron variant (Chung et al., 2021; Xia et al., 2022).

Remarkably, it is estimated that COVID-19 vaccinations prevented more than 14 million deaths in only one year after global vaccination enrollment^4^. This was accomplished by breakthrough innovations in vaccinology, in addition to global efforts to fund worldwide clinical trials, including classical and new-generation vaccines^5^. The mRNA vaccines (RNA) Comirnaty® (BNT162b2, BNT) and Spikevax® (mRNA-1273, MO), the viral vectored (VV) vaccines Covishield®/Vaxzebria® (AZD1222, ChAdOx-1, ChAd) and Jcovden® (Janssen), and inactivated vaccines (IN) such as Coronavac® and Covilo® (BBIBP-CORV, BBIBP) were among the first and most used vaccines worldwide until 2023 ^6^. Particularly, ChAd and BNT were the most used vaccines during the first years of vaccination and 2 and 3-dose heterologous regimens were widely used and assessed ^7,8^. The combination of different vaccines and platforms shows how classical and new-generation vaccines can help control epidemics and potentially trigger complementary systemic immune responses that individual vaccine technologies are intrinsically incapable of ^8–11^. The complete homologous vaccination regimen showed high efficacy in clinical trials against COVID-19, as high as 95.0%, 94.1%, and 90% for new-generation vaccines, such as BNT, MO, and ChAd, respectively ^11–13^. Classical technologies, such as inactivated virus (IN), and protein subunit (SU) vaccines, have also shown high efficacy and effectiveness. BBIBP and Zifivax® (ZF2001) had 78.1% and 75.7% efficacy against symptomatic infections, respectively ^14,15^. Although these vaccines were initially tested and approved in homologous regimens, heterologous vaccination has been tested in clinical trials and real-world scenarios to address ongoing waves of infection and new VOCs ^8,16^.

To understand how different vaccination regimens and vaccine technologies affect the immune system in healthy individuals and infected patients during, and after infection by SARS-CoV-2, it was integrated RNAseq data from five distinct studies that include vaccinations with Covilo®, Vaxzebria® or Covishield®, Spikevax®, Comirnaty®, and Zifivax®. These studies, however, present data from different time points, populations and regimens, and there is a need to obtain a unified perspective of their immunological signatures. This study introduces a systems vaccinology atlas that delineates shared and individual profiles across conditions in both healthy and infected individuals. The findings indicate that completing a vaccination regimen can effectively limit disease progression. Additionally, it demonstrates that heterologous vaccination strategies can diversify immunological signatures, potentially enhancing protection against the disease. Furthermore, the study explores immune responses induced by vaccines targeting other pathogens and reveals a substantial overlap in genes related to the innate immune system with COVID-19 vaccines.

## 2. Results

### Immune-related gene expression varies by vaccine type, regimen, and infection status

A total of 562 samples were analyzed, sourced from peripheral blood mononuclear cells (PBMCs) of 245 participants involved in five distinct studies (**Table 1**, **Fig. 1A**). These participants were either vaccinated (V) with one or more of the four different types of vaccines (IN, SU, RNA, and VV), which comprised both homologous (HO) and heterologous (HE) vaccination regimens and were administered in one (V1), two (V2), or three doses (V3), or were infected patients (I) with or without prior vaccination (**Fig. 1B-C, Fig. S1-2**). Many issues were identified with the GSE206023 count matrix, in which most of the genes were annotated in GenBank IDs with multiple identifier versions for the same genes. Moreover, only 10% of these identifiers could be annotated, comprising less than 2% of the immune genes, in contrast to more than 90% present in the other datasets. Given the importance of this dataset regarding the diversity of vaccine technologies, a decision was made to reprocess the read files in FASTQ and align it with the reference transcriptome using pseudoalignment in the Kallisto package ^17^. While other datasets employed HISAT2, pseudoalignment was chosen due to its superior speed, robustness, and lower computational resource requirements compared to HISAT2. ^18^.

**Fig. 1.**
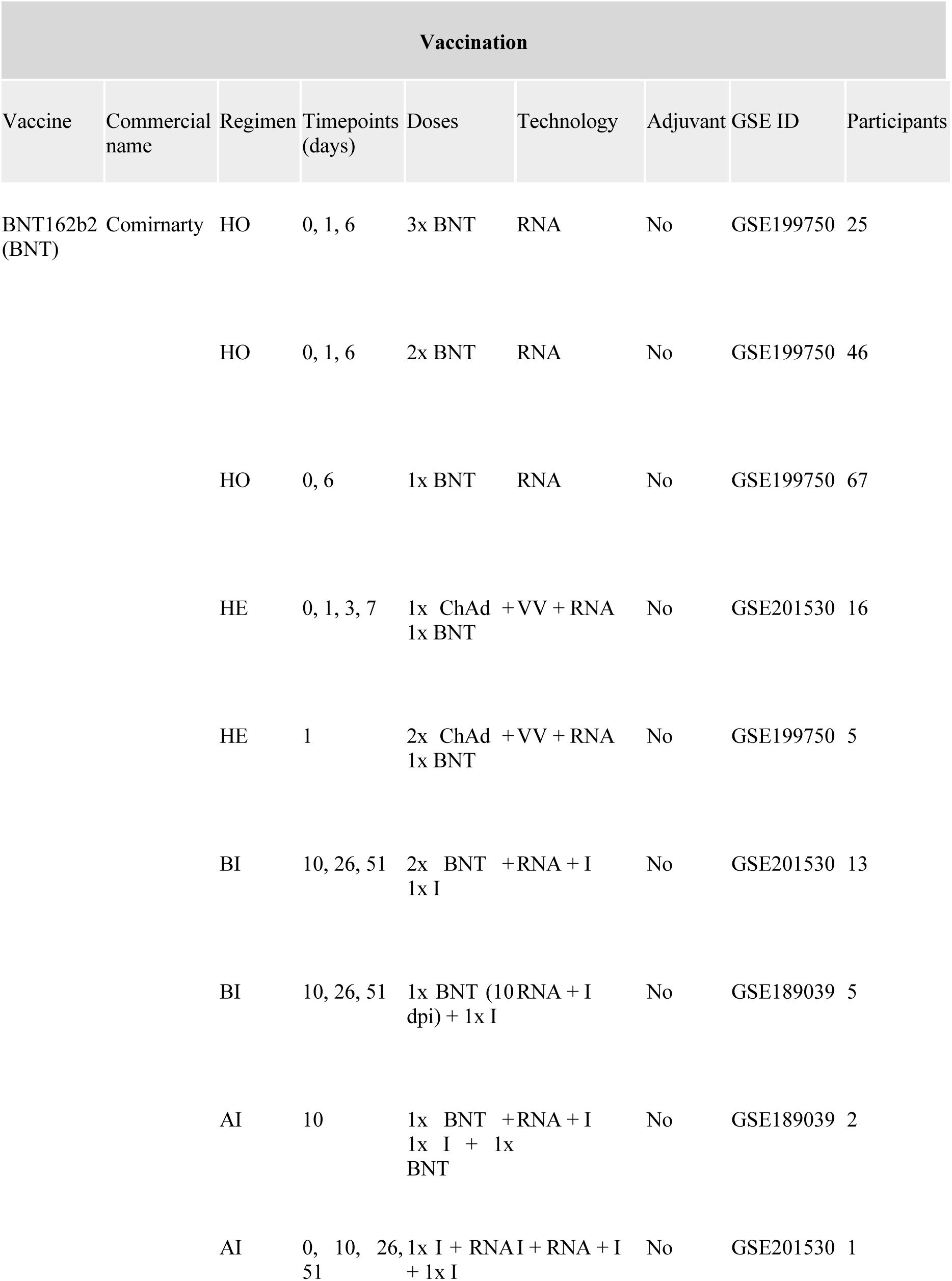
Methods and description of datasets and study population. (A) We manually curated the GEO datasets and performed differential gene expression analysis. Finally, genes from immune system biological process (GO:) and the VAX collection from MSigDB were used in GSEA and ORA, and individual gene expression analysis was analyzed across the studied conditions. Created with Biorender. Number of participants (B) and samples (C) were categorized by vaccine and infection conditions. Legend: BBIBP: BBIBP-CORV; BNT: BNT162b2, ChAd: ChAdOx-1; MO: mRNA-1273; I: Infected; V: Vaccinated.

**Table 1.**
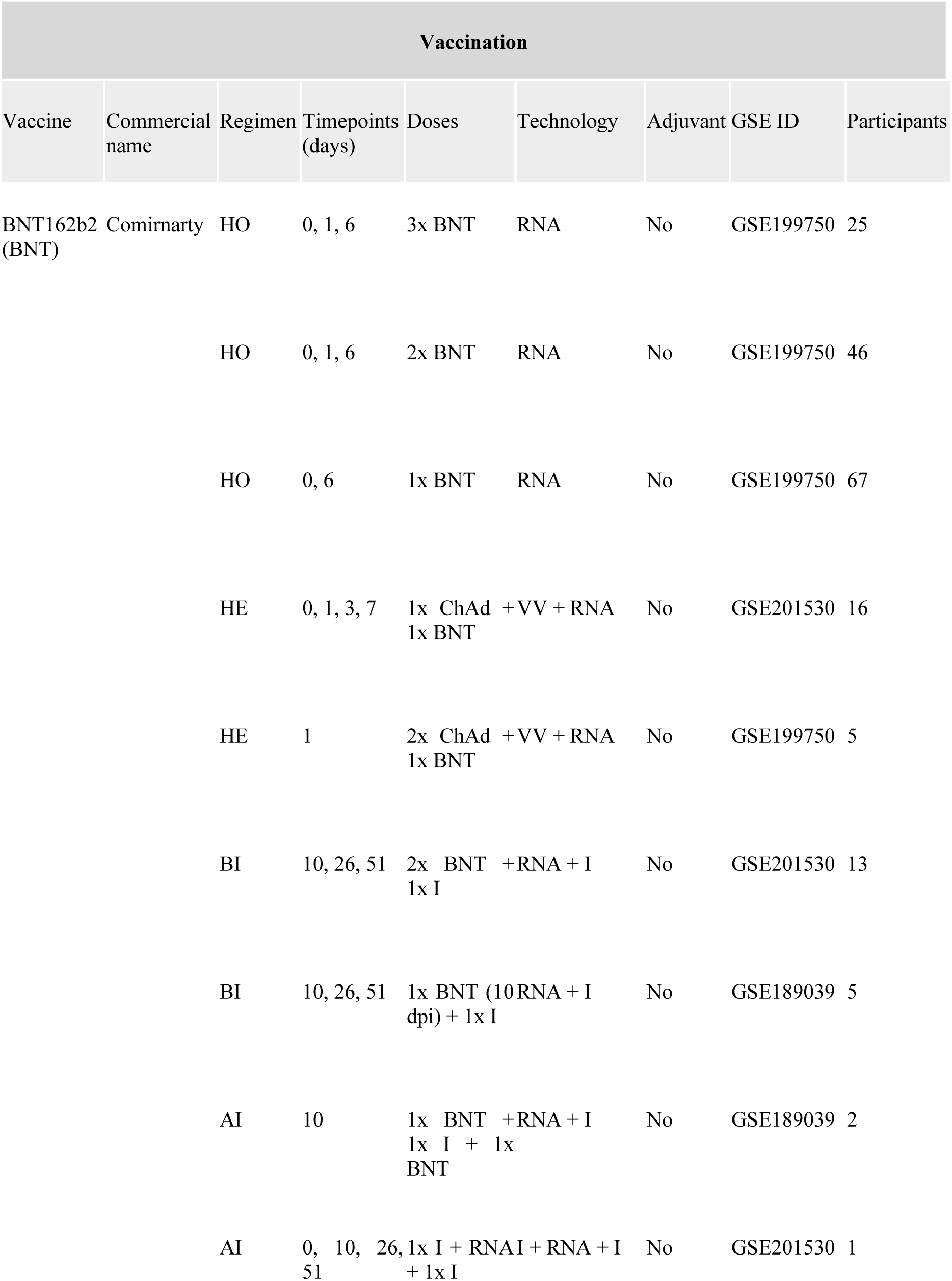
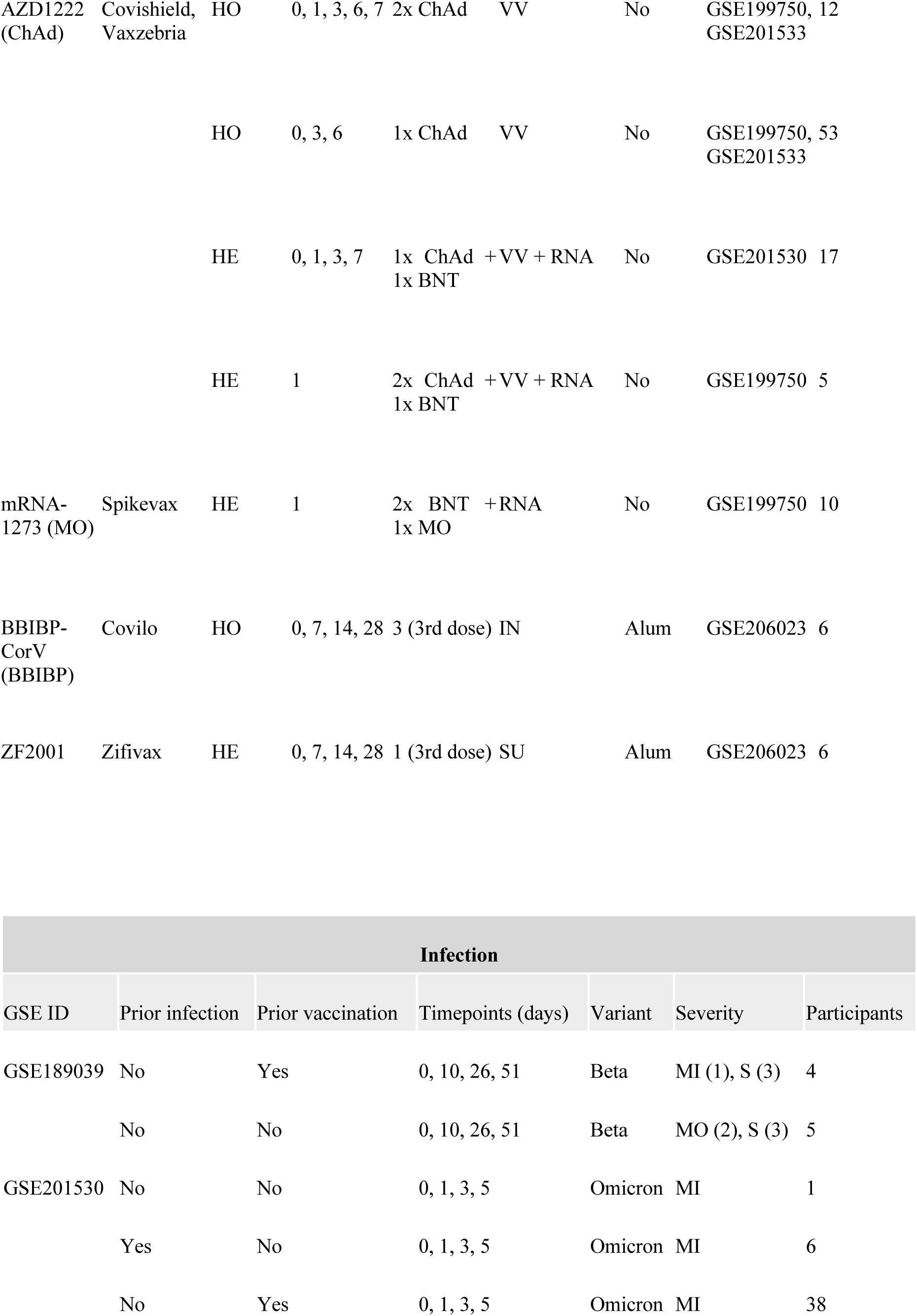
Vaccines and infection conditions selected in this study. Legend: HO: Homologous; HE: Heterologous; BI: Before infection; AI: After infection; I: Infection; RNA: mRNA; VV: Viral vector; IN: Inactivated; SU: Subunit. MI: Mild; MO: Moderate; S: Severe; A: Asymptomatic.

The temporal dynamics of differentially expressed genes (DEGs) was influenced by vaccine type, number of doses, and administration regimen. This happened across convalescent and non-convalescent individuals, as well as infected patients with previous vaccination (**Fig. 2A, Fig. S3**). Notably, the BNT vaccine exhibited a significant increase in total DEGs across the three doses, with three DEGs six days after the first dose (BNT V1), followed by 613 and 96 DEGs 1 day after the second (BNT V2) and third doses (BNT V3), respectively, with a more substantial increase with two doses of BNT followed by a single dose of MO (BNT-MO V3) (**Fig. 2A)**. In contrast, it was observed that the ChAd vaccine exhibited a divergent pattern compared to BNT in non-convalescent individuals, with decreased up– and downregulated DEGs over the first week after two homologous doses (ChAd V2), and a higher number of DEGs in the heterologous vaccinations (ChAd-BNT V2 and ChAd-BNT V3) (**Fig. 2A, Fig. S3**). A similar trend was observed in homologous and heterologous vaccinations with the BBIBP vaccine after a third dose (**Fig. 2A, Fig. S3**). It was evident that homologous vaccination with an inactivated virus vaccine does not generate significant DEGs in a third dose (BBIBP V3) and thus requires heterologous vaccination with a different technology, such as a SU vaccine (ZF2001 V3) (**Fig. 2A, Fig. S3)**.

**Fig. 2.**
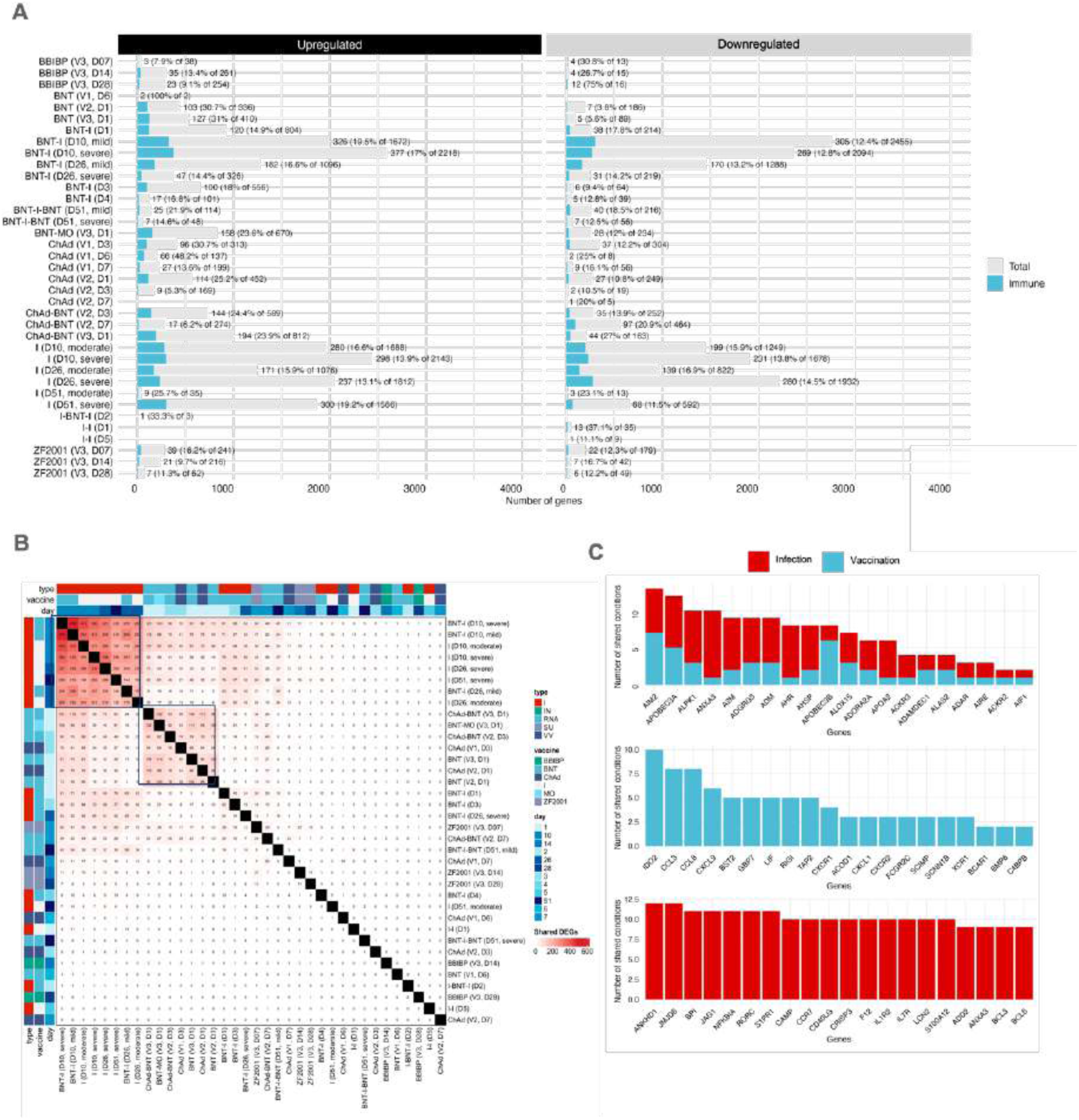
A. Differentially expressed genes (DEGs) were categorized as up-(left panel) and down-regulated (right panel) with the absolute and relative number of genes. The conditions were then plotted with the total number of genes and immune genes. Labels correspond to the total number of immune genes and the percentage over total DEGs. B. Shared DEGs between infection and vaccination. The upper-left cluster include individuals infected for the first time, and the bottom-right cluster comprise the first week of vaccination by RNA, VV, and SU vaccines. C. Twenty most shared immune-related genes. Legend: BBIBP: BBIBP-CORV; BNT: BNT162b2, ChAd: ChAdOx-1; MO: mRNA-1273; I: Infected; V: Vaccinated; IN: Inactivated; RNA: mRNA vaccines; SU: Subunit; VV: Viral vector. B. Shared DEGs in vaccination and infection.

Furthermore, infections in vaccinated and unvaccinated patients notably affected the number of downregulated DEGs (**Fig. 2A, Fig. S3**). Among unvaccinated infected individuals, DEGs were higher in severe disease cases than in moderate cases over 51 days. Conversely, in infected patients with prior complete vaccination, primarily mild and moderate disease, this number was at least 10 times lower. However, infected hospitalized patients with moderate and severe disease who were vaccinated with a single dose of BNT 10-11 days before hospitalization, the number of DEGs was similar to that of infected patients on days 10 and 26. This pattern could also be influenced by the age range (71-90 years) of these patients, which is a major risk factor for COVID-19^19^. Of the four infected patients, three recovered, and one died (**Fig. S3)**. The surviving patients continued dexamethasone treatment and received a second dose of the vaccine 40 days post-infection^20^. Samples were collected 15-20 days post-vaccination, exhibiting more DEGs than at a later time post-vaccination in non-convalescent individuals with BNT or ChAd (**Fig. 2A)**. The gene expression patterns in unvaccinated and single-dose vaccinated patients during the second infection were similar, but vaccinated individuals exhibited a notable, threefold reduction in DEGs compared to with unvaccinated counterparts. Only in the total DEGs analysis, without discriminating by ontology (immune and non-immune), was an apparent relationship between vaccine type, disease severity, and time.

Upon filtering immune-related DEGs, it became clear that the fraction of immune-related genes decreased over time in vaccinees and infected patients (**Fig. 2A, Fig. S3**). Specifically, this investigation focused solely the immune-related genes excluding genes from the BCR and TCR repertoire [immunoglobulin chains (IG@) and T-cell receptors (TR@)]. Particularly in infected patients, the first days showed a greater fraction of immune genes, which decreased over time, whereas the number of other non-immune-related genes increased (**Fig. 2A, Fig. S3A**). In addition, infected patients with prior complete immunization showed fewer DEGs and a smaller fraction of immune genes. Nevertheless, vaccination results in the activation substantially more immune genes, which follow proportionally to other genes, either increasing or decreasing, depending on the vaccine type. This contrast indicates that the immune response induced by vaccines is quantitatively more balanced in terms of the expression of immune genes than infection. To identify patterns among these conditions, immune-related and non-immune DEGs were compared distinctively (q-value < 0.10) (**Fig. 2B, Fig. S4A**). It was observed that the majority of shared genes among the different vaccine types were non-immune genes, such as ANKRD22, GBP1P1, GPRB4, and HES4 (**Fig. S4A-B).** Moreover, the most shared genes, such as LRRN3, ADAMTS2, and FAM209B, were more present among infected patients (**Fig. 5B**). This shows the role of other processes in vaccination and, to a greater extent, infection.

To understand these non-immune processes during vaccination, the analysis focused on the non-immune genes. It was found that a majority of these genes were related to ubiquitous cellular and metabolic processes, and the regulation of biological processes (**Fig. 2A-B, Fig. S4**). However, the subsequent analysis specifically targeted immune genes. The shared immune DEGs exhibited one central cluster associated with the groups infected for the first time (vaccinated and unvaccinated) and another cluster comprising the first day of vaccination with homologous or heterologous second and third doses of ChAd, BNT, and MO (**Fig. 2B**). Notably, the number of shared genes among infected groups was 2– to 10-fold higher than non-convalescent-vaccinated groups. In addition, it was observed a decrease in shared DEGs between infected patients with prior vaccination (BNT-I) and infected patients with prior infection and prior vaccination (I-BNT-I), in a similar proportion to non-convalescent vaccinated groups, which was not evident in the non-immune DEGs context (**Fig. 3B, Fig. S4A**). This finding was also associated with vaccine type, viral vector vaccines (ChAd and ChAd-BNT), and subunit vaccines (ZF2001), which were more evident with non-immune DEGs (**Fig. S4A)**.

**Fig. 3.**
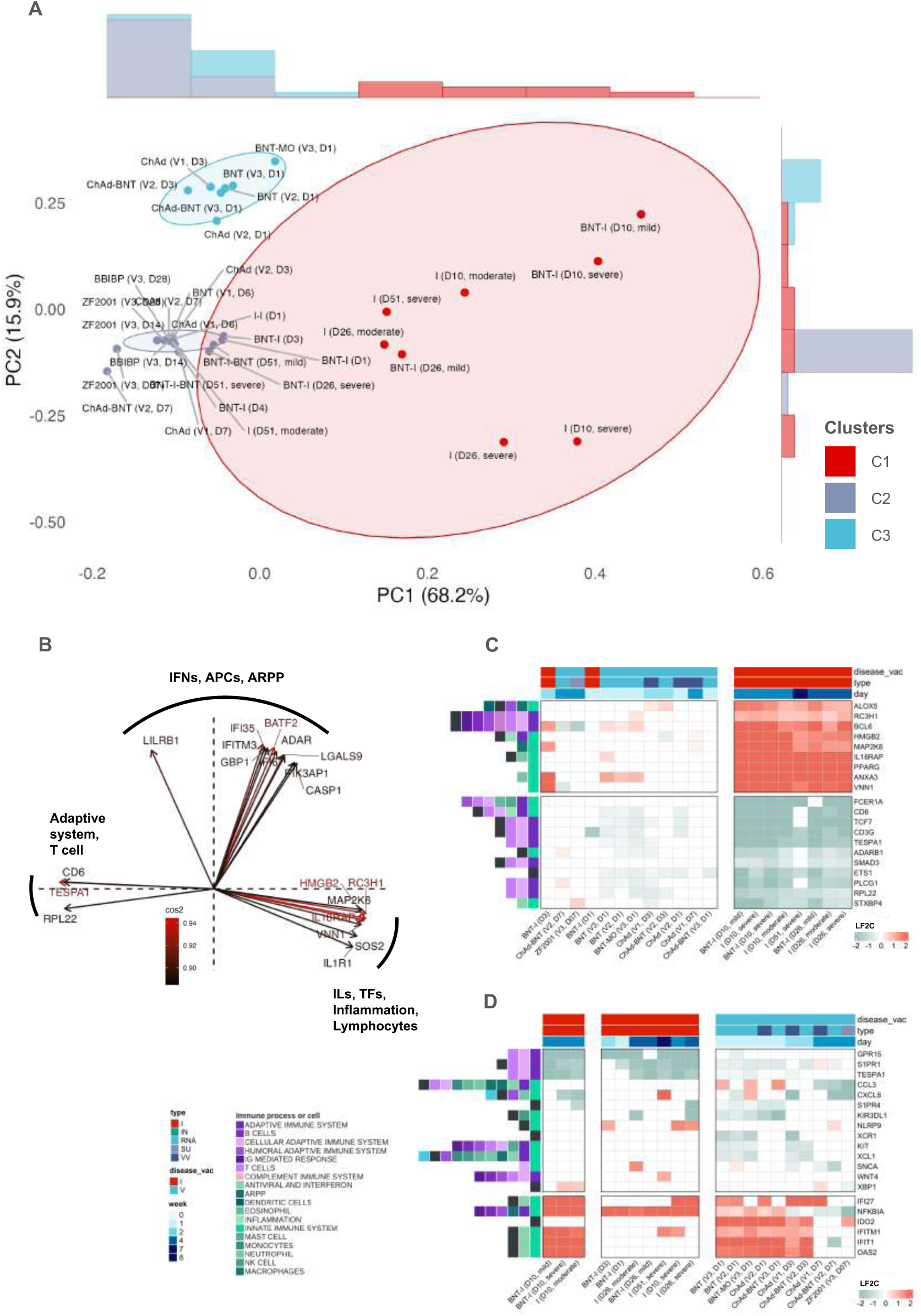
Principal component analysis. A. Two principal components were employed to plot all conditions, capturing 86.2% of the total variance. K-nearest neighbors (KNN) clustering identified three distinct clusters. Cluster 1 (C1) includes infected patients and overlaps with Cluster 2 (C2). C2 is predominantly characterized by vaccination in healthy individuals, encompassing convalescent patients and non-convalescent individuals. Cluster 3 (C3) comprises the initial three days post-vaccination with mRNA vaccines and ChAd. B. The loading plot displays cos2 values for the top 20 genes in PC1 and PC2. The top 20 genes contributing to PC1 (C) and PC2 (D), along with the corresponding Log2 Fold Change (L2FC). Genes were displayed with correspondent log2-fold change ranging from –2 (blue) to 2 (red). The left annotation on the heatmap depicts the contribution to different immune processes and cells, with colors corresponding to processes and cells related to the complement system (pink), the innate (greens) and adaptive immune systems (purples), and general processes and cells (leukocytes, black). Legend: BBIBP: BBIBP-CORV; BNT: BNT162b2, ChAd: ChAdOx-1; MO: mRNA-1273; I: Infected; V: Vaccinated.

In contrast, there was a noticeable decrease in the number of genes on subsequent days, particularly on day 7, suggesting the initiation of homeostatic processes over time. This reduction in DEGs becomes more pronounced on subsequent days. However, this decrease contrasts with the trend observed with increasing doses, particularly notable in homologous RNA vaccines and heterologous ChAd with subsequent vaccination with BNT. This indicates a broader activation of diverse genes with a higher number of doses. Particularly, vaccination with BNT following one or two doses of ChAd led to a significantly greater number of total DEGs compared to the two-dose homologous regimen of either vaccine (**Fig. 2A-B, Fig. S4**). Comparing to the two doses of ChAd on day 3, there was a three-fold increase in upregulated DEGs, which reduced by half on day 7. Conversely, downregulated DEGs were 13-fold higher in the ChAd-BNT regimen but increased two-fold on day 7. Remarkably, even though the assessment was conducted only one day post-vaccination, upregulated DEGs exhibited in the ChAd-ChAd-BNT regimen were approximately two-fold higher than those with two doses of ChAd and 1.2-fold higher than those with BNT-MO.

In contrast, the number of up– and downregulated genes was nearly equivalent in infected individuals, regardless of prior vaccination status (**Fig. 2A, Fig. S4A**). Additionally, genes from unvaccinated infected individuals were largely constrained within their group and had a limited overlap with vaccinated individuals.

To describe the immune transcriptome of individual vaccination regimens, were compared all the genes present in the datasets with the immune system related genes (**Fig. 4D**). Following this comparison, the enrichment of gene sets associated with immune responses was analyzed. (**Fig. 4C**).

**Fig. 4.**
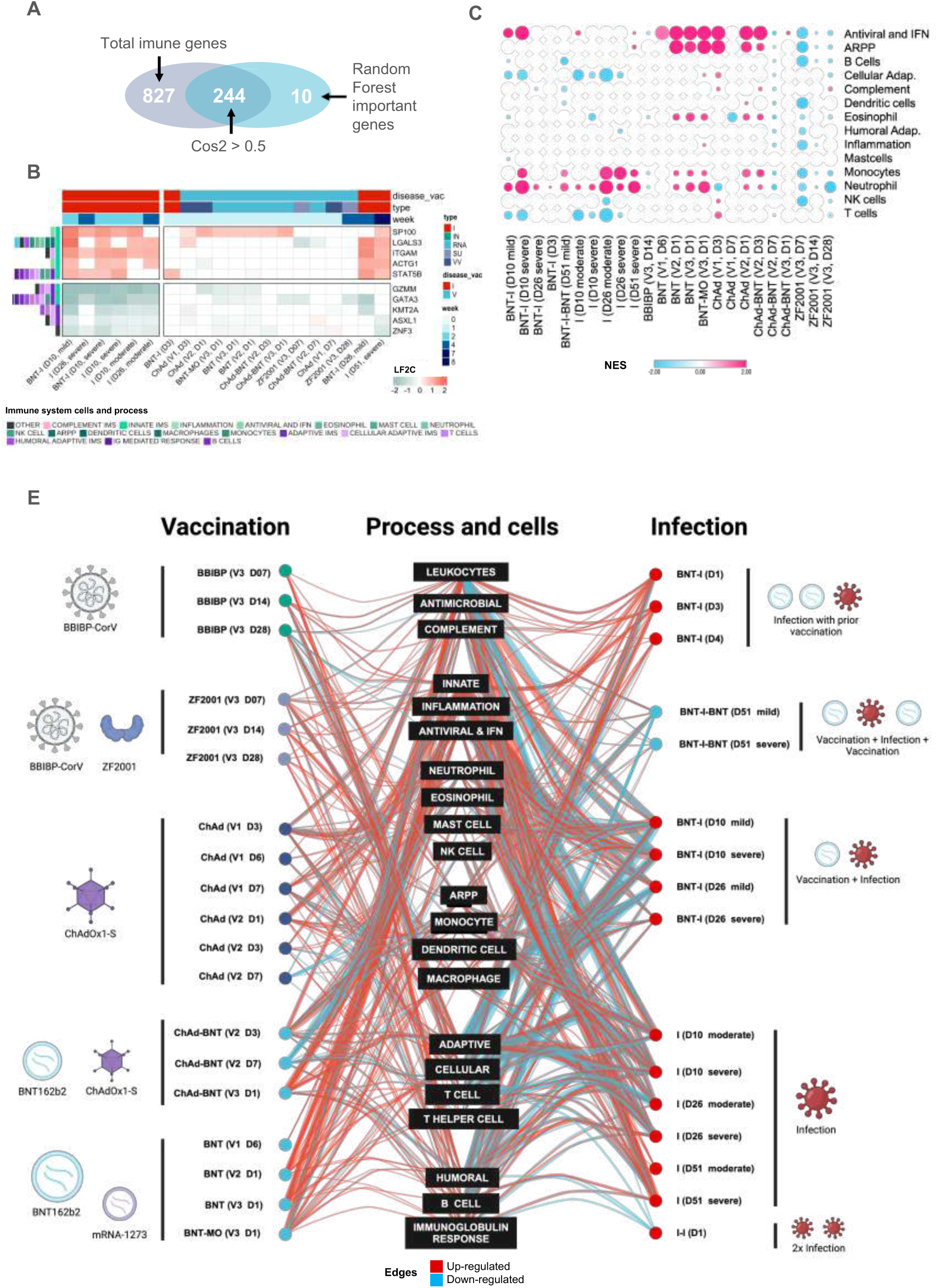
DEGs with cos2 > 0.5 were used to train a random forest model to predict the types of vaccines and infection, and the ten most important genes were selected (A). The final model was selected after an iterative process of hyperparameter tuning. The best model was defined with 2000 trees in the forest, where mtry was set to 6 and min_n to 6. The most important genes were used to plot the log2-fold change (L2FC) (B). C. Gene set enrichment analysis of COVID-19 vaccines and infection. Major immunological gene sets were enriched with GSEA. Normalized enrichment scores (NES) for enriched gene sets, with circle size corresponding to –log(qvalue). D. Representation of upregulated (red) and downregulated (blue) genes in COVID-19 conditions associated with various immune biological processes. Visualization IN stacked hierarchical layout was generated using Cytoscape ^61^. Node size is determined by degree centrality, and node colors are determined by type and infection status. Illustration made in **BioRender.** Legend: BBIBP: BBIBP-CORV; BNT: BNT162b2, ChAd: ChAdOx-1; MO: mRNA-1273; I: Infected; V: Vaccinated.

**Fig. 5.**
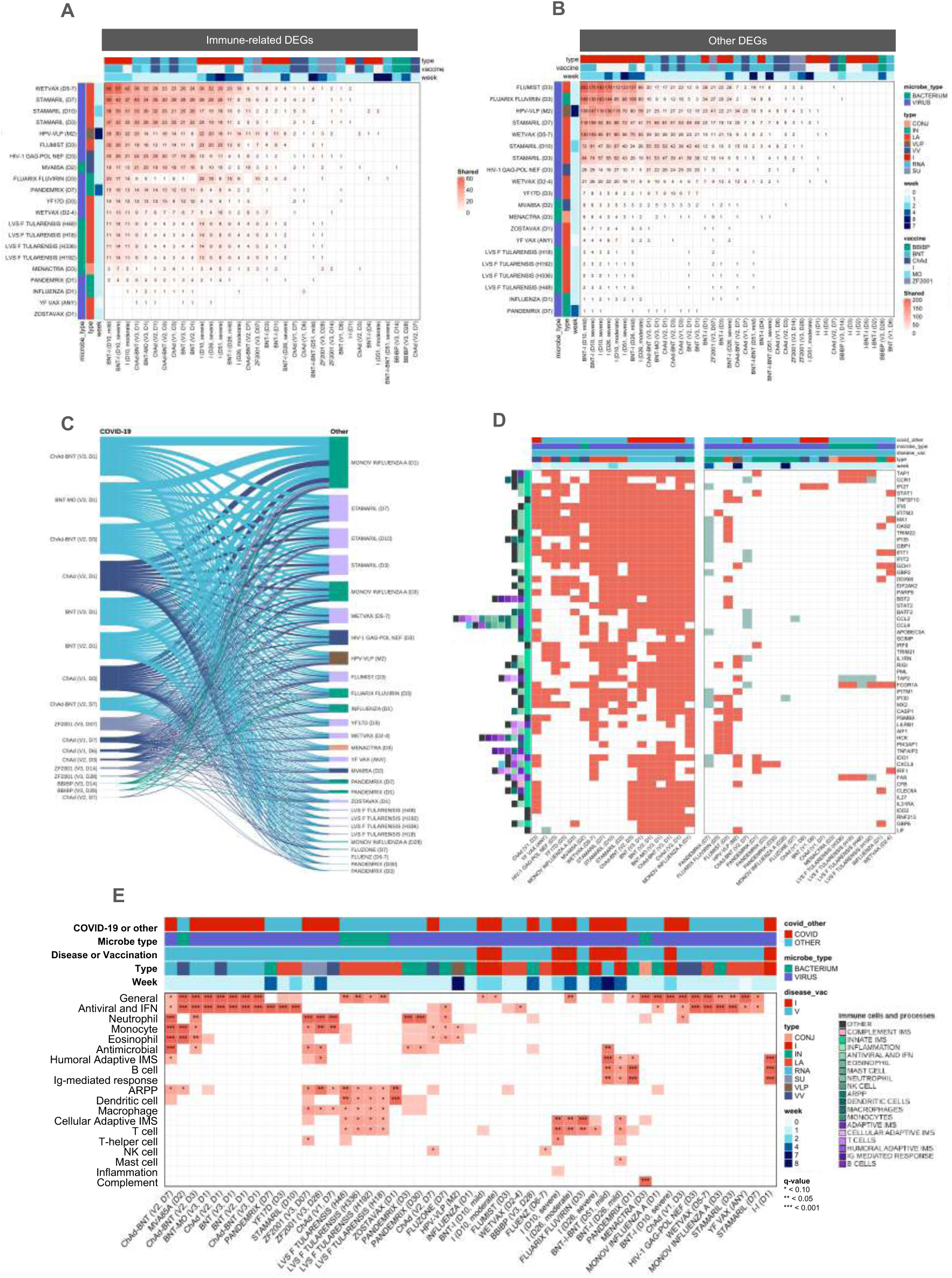
Gene-level comparison of COVID-19 vaccines and the MSigDB Vax collection. Overlapping immune-related (A) and other (B) genes between conditions. C. Genes with the most sharing between COVID-19 vaccination and other vaccines. Visualization in stacked hierarchical layout was generated using Cytoscape ^61^. C. All immune-related genes shared at least once between conditions. D. Genes included in the first cluster (see complete heatmap in **Fig. S9**) were set with L2FC as 1 (upregulated) and –1 (downregulated). These genes were annotated by their corresponding contribution to subprocesses and cells among the innate, adaptive and complement immune systems (left). E. Over-representation analysis of vaccines against COVID-19 and other pathogens with immune cells and processes. Legend: BBIBP: BBIBP-CORV; BNT: BNT162b2, ChAd: ChAdOx-1; MO: mRNA-1273; I: Infected; V: Vaccinated.

### Limited Immunological Profile Observed with Three Homologous doses of BBIBP

The BBIBP exhibited enrichment of processes associated with the innate immune system, including the antiviral response mediated by interferons, neutrophils, eosinophils, and monocytes, which were consistently found to be negatively regulated (**Fig. 4C**). Further analysis of gene expression within these processes and cells revealed specific genes of interest (**Fig. S4-7)**. On day 14, we identified genes associated with inflammation (NLRP9, upregulated), immune regulation (IDO2), macrophage-mediated antiviral response (CCL3, downregulated), neutrophil activation (CXCL8), and lymphocyte-macrophage signaling (WNT4, upregulated)^21^. On day 28, a transcription factor (NFKBIA, downregulated) present in ARPP and B cells, which regulate immune system processes, and CCL2 (downregulated), a monocyte chemokine, were observed.

### ZF2001 as Heterologous Booster Elicits Distinct Immune Response Profile

A heterologous regimen with the ZF2001 vaccine in individuals who had previously received two doses of the BBIBP vaccine resulted in a distinct gene expression profile compared to the homologous vaccine. Although most DEGs were upregulated, a small fraction of genes related to the immune system were downregulated (**Fig. 2A**). On the seventh day post-vaccination, there was a suppression of genes associated with inflammatory processes (IL1RN, FOSL2, and CD58), antiviral response and interferons (CXCL2, CXCL3, CCL3, CCL4, CCL20, BCL3, ADM, LGALS3, FAU), as well as genes related to antigen recognition and presentation (TNFAIP3, FOSL1, RAB3B, and ICAM). This suppression also extended to genes expressed in eosinophils (VAMP2), NK cells (RNF19B), dendritic cells (NFKBIA, FOSL1), T cells (IL2), B cells, and immunoglobulin-mediated responses (BCL3, BCL6) **(Fig. 2C**, **Fig. S6-9**).

On the other hand, a predominance of upregulated genes was observed on the seventh day, particularly associated with other chemokines (CX3CL1, CXCL8, CXCL14, CCL8, CX3CL1), mast cells (CPLX2, CLNK, LGALS3), ARPP (ITGB8, TRIL), APCs (ADGRF5, FLT1, IL33, WNT4), adaptive response processes and cells (CACNB4, EOMES, SPNS2, BCAR1), as well as the complement system (**Fig. S6-9)**. Specifically, within the complement system, genes from the classical pathway (C4B and C4BPB) and lectins (MASP1 and MASP2) were overexpressed (**Fig. S6-9)**. In the adaptive immune system, there were genes related to T cell activation (IL13, EPHB1, TNFAIP8L2, TARM1), specifically Th2 response (IL33), Th17 (CCR2), TCD8 (EMP2), B cell and Ig-mediated response (WNT3A) (**Fig. S6-9)**.

### Viral Vectored Vaccine ChAd Induces Early Gene Expression of Complement and Adaptive Immune Systems

Genes related to T cells and the cellular response to the viral vector were expressed as early as the third-day post-vaccination with the first dose. Notably, genes such as CCNB2 and CAV1, which play roles in the cell cycle and T cell activation, respectively, exhibited sustained expression levels up to the seventh day. Additionally, the innate immune system processes are enriched, including interferon-mediated antiviral responses, acute phase response proteins (ARPP), and NK cell, related proteins along with upregulated genes from the complement system associated with the alternative pathway (CFB and CFD) (**Fig. S7-9**).

It is also essential to highlight the distinct expression of genes related to humoral responses on the third and seventh days (**Fig. S8**). These expression patterns were also observed in B-cell genes, with overexpression between days 3, 6, and 7, returning to the initial profile by the sixth day. Conversely, except for genes associated to cellular response, B cells processes, and the humoral immune response were enriched on the first day after the second dose. However, gene-level analysis still revealed diverse and high gene expression at this stage, especially for the cytokine CCL2 and immunoregulatory enzyme IDO1. When comparing genes related to the humoral response between doses, it was noted that these genes were less expressed and less diverse at the second dose. On the third day, only two DEGs associated with the humoral response were identified (SYK and KIT). Particularly, on day 7, CD27, a gene involved in immunoglobulin synthesis, ^22^ was increased after the first dose and suppressed after the second dose.

### Distinct Gene Expression Patterns Elicited by mRNA Vaccines BNT and MO

The mRNA vaccines BNT and MO demonstrate distinct gene expression patterns. Six days after the first dose of BNT, only 3 DEGs were identified, with IFI27 being the only immune-related gene (**Fig. 2A, Fig. S4-7)**. Subsequent second and third BNT doses revealed elevated activation of antiviral and interferon responses, ARPP, and the function of neutrophils, eosinophils, monocytes and dendritic cells (**Fig. 4C**). These sets were similarly enriched following the third homologous vaccination, except for dendritic cells. A third heterologous vaccination with MO induced dendritic cell function, inflammation, and the activation of the cellular adaptive immune system. Notably, IFI27 expression was exclusive to all BNT doses except for the third dose of MO (**Fig. S4-7)**. Neutrophils shared genes in various vaccination regimens, including CCL8, CXCL9, and CCL2 (**Fig. S4-7)** ^23^. In addition, eosinophils exhibited pronounced CCL3L1 expression following MO vaccination, which plays a pivotal role in virus clearance from the lungs ^24^.

### Immune Signatures in Infected and Convalescent Patients Before and After Vaccination with BNT

To understand the impact of vaccination before and after infection, we analyzed infected and convalescent patients. We found that innate immune cells and processes were highly activated and persisted for several days, especially interferons 1 and 2, neutrophils, APCs, and bacterium-directed PRRs (TLR4 and NOD) **(Fig. 4C, Fig. S4-7).** Additionally, B-cell processes, including chemotaxis and homeostasis, are activated **(Fig. 4C, Fig. S4-7).** There was also a Th2 response in BNT-I (D26, mild), although T cell processes were predominantly downregulated under other infection conditions. In a gene-level analysis, many genes related to the adaptive immune system were upregulated and downregulated. Likewise, it was evident that infection induced different subsets of genes compared to vaccination in every immune process and cell type (**Fig. S4-7)**. Considering the innate immune system and classical pathway complement system genes (such as CR1 and C1R), high and persistent gene expression, especially those associated with inflammation (e.g., IL10, HMGB2, ELANE, and NLRP6), activation of neutrophils (e.g., CD177, ANXA3, and IL18RAP) and mast cells (e.g., S100A12 and SLC15A4), may be associated with the initiation of cytokine storms, allergic reactions, and thrombosis ^25,26^. Although most genes participating in adaptive immune system processes were highly downregulated, there were also many highly expressed genes in the humoral response, including genes related to Ig-mediated responses, ARPP, and other B-cell processes, as well as in cellular response and T-cell processes. This pattern varied among convalescent and infected patients who had received prior vaccination, regardless of whether they had a completed or incomplete vaccine regimen (**Fig. S4**).

### Stratification of COVID-19 vaccination and infection using multivariate analyses

To describe the similarities and differences among all conditions, it was conducted a Principal components analysis (PCA) with all immune DEGs (**Fig. 3A-D**). Principal component 1 (PC1) accounted for 66.8% of the variance, whereas PC2 accounted for 15.7%, totaling 81.9%. PC1 was predominantly defined by genes associated with interleukin (IL) signaling pathways (IL18RAP, SOS2, MAP2K6), apoptosis (HMGB2), inflammation-related processes (RC3H1, ALOX5, PPARG, and FCER1A), as well as processes linked to lymphocytes (ETS1), particularly to T cells (VNN1, CD6, TESPA1, RPL22, PCLG1, TCF7, TRAT1, and CD3G) **(Fig. 3C)**. On the other hand, PC2 was defined by genes related to antiviral and interferon responses (IFI35, IFITM3, STAT1-2, IDO1, ADAR, BST2, GBP1, and PML), as well as APCs (BATF2, LILRB1) and antigen receptors, processing and presentation (ARPP) (TAP2, PDIA3, TRIM26, and CTSS) **(Fig. 3D).**

The results revealed the presence of three primary clusters (**Fig. 3A**). The first significant cluster indicated patients with ongoing infection, characterized by genes associated with IL-signaling pathways, apoptosis, and inflammation-related processes in cluster 1 (**C1; Fig. 3A,C).** While this cluster included specific individuals with previous vaccination, we did not anticipate a substantial influence of vaccination, particularly considering that it was administered 10 days before the onset of COVID-19 symptoms. Nevertheless, it is evident that patients who received a complete vaccination regimen and were subsequently infected (BNT-I), as well as a patient who received a second dose of BNT 10 days after recovery (BNT-I-BNT, D51), showed overlap with Cluster 2 (C2). The C2 cluster comprises homologous vaccinations for both convalescent patients and non-convalescent individuals. Additionally, healthy-vaccinated individuals were part of this cluster, but it also included patients with two infections (without severe outcomes) and one patient initially experiencing severe disease 51 days after the onset of symptoms, who were hospitalized and treated with steroids. Given that this cluster occupies the central position in the plot and represents comparatively healthy conditions, the proximity of eigenvalues from the first cluster to the second cluster suggests an ongoing, time-dependent homeostatic process. In contrast, the third cluster (C3) consisted of homologous and heterologous vaccinations involving genetic-based vaccines such as BNT, ChAd, and MO, specifically within the initial three days post-vaccination. This cluster is characterized by genes in PC2 and shares a minimum number of certain genes with infected patients in C1. Nevertheless, the expression of the top 20 contributing genes in PC2 was significantly higher in C3 than in C1 conditions (**Fig. 3B,D**). These differences were marked by genes such as IDO1, IL31RA, and LILRB1, which were not expressed during infection and are associated with antimicrobial response, chemotaxis, and MHC1 presentation, respectively.

### Machine learning classification of COVID-19 vaccine types and infection based on immune genes

Using the PCA results, genes in PC1 and PC2 (cos2 > 0.5) were filtered and a random forest model was trained for classification based on the variable “type”, including infection and the different vaccine types (**Fig. 4A**). This model demonstrated remarkable performance, with an accuracy of 91.2%, an area under the receiver-operator curve (ROC) of 0.990, and an out-of-bag (OOB) error of 0.086. The main gene predictors were associated with transcriptional regulation (GATA3, ZNF3, KMT2A, and ASXL1), antiviral response (SP100), granzyme (GZMM), ARPP (ITGAM, ACTG1, LGALS3), and T cell response (STAT5B) (**Fig. 4B**). The high expression of the transcriptional regulation associated genes GATA3, ZNF3, KMT2A, and ASXL1, an antiviral response gene (SP100), and the granzyme GZMM, were key markers for infection, along with the suppression of genes that play roles in ARPP (ITGAM, ACTG1, LGALS3), and T-cell response (STAT5B) SP100 was also notably expressed in vaccination, in the first 3 days after the second and third doses of both homologous and heterologous regimens. LGALS3 was also markedly expressed in the first 3 days of vaccination with ChAd. This observation aligns with the fact that viral vectored vaccines, like ChAd, act through certain infection associated mechanisms to deliver genetic material into the nucleus^27^.

### Comparative Analysis of Vaccines Against Other Pathogens

Understanding how COVID-19 vaccines relate to classical and consolidated vaccines used in routine immunization is essential. Therefore, all immune-related DEGs associated with various vaccines, including COVID-19, were compared using MSigDB Vax collection^28^. The dataset was manually annotated and selected all up– and downregulated genes from vaccination-only sets (**see Methods, Fig. 1A**). This approach originated from several gene sets, including those correlated with antibody titers, T-cell responses, and training sets. To understand the extent of gene overlap, was performed a gene-level analysis of these shared genes from PBMC-derived samples, to match the COVID-19 conditions, while filtering DEGs with –1 < L2FC > 1 (**Fig. 5A-C, Fig. S8 and 9**).

The analysis uncovered a notable overlap of genes, both up– and downregulated, between vaccines targeting viruses and bacteria based on inactivated (IN) and live-attenuated (LA) pathogens and COVID-19 vaccines (**Fig. 5A-C**). This overlap was most prominent in the initial two weeks and following the second and third doses. Specifically, downregulated DEGs were predominantly associated with infection, particularly with LA vaccines (**Fig. 5A**). Conversely, upregulated DEGs were common among vaccinated individuals, comprising both non-convalescent and convalescent individuals. This overlap extended to include infected patients who had undergone prior one-dose vaccination.

The observations revealed two main clusters of genes, predominantly representing the innate and adaptive immune systems (**Fig. S6)**. While the second cluster encompasses genes present in the adaptive immune system, these genes also exhibit a lower degree of overlap with the innate immune system. Notably, the second cluster appeared more heterogeneous and demonstrated significantly fewer shared genes across vaccines than the first cluster. In this first cluster, we observed that COVID-19 vaccines and others shared most innate genes that also play a role in the adaptive immune system (**Fig. 5D**). Some of the most common genes between vaccines included chemokines and receptors (CCL2, CCL3, CCL8, CCL20, CCR1, and CXCL8), interferons and antiviral responses (IRF1, IFIT1, STAT1, and STAT2), ARPP (TAP1, TAP2, HLA-DMB, and HLA-DMA), and some adaptive immune system genes (RIGI, FAS, CD83, BCL6, and FCER1G) (**Fig. 5B**). On the other hand, some of the genes present mainly in COVID-19 vaccine conditions are chemokines, such as CCL2, which play a diverse role in the innate, adaptive, and complement systems (**Fig. 5C**). CCL2 is also shared with the monovalent influenza vaccine adjuvanted with AS03 and the LA vaccine Fluzone®^29^. Although most non-COVID-19 vaccines sharing genes with COVID-19 vaccines are targeted against the influenza virus, Stamaril®, a yellow fever LA vaccine, shares many innate immune system genes with COVID-19 vaccines ^30^. Indeed, an overrepresentation analysis (ORA, FDR < 0.25) of immune genes showed that general leukocyte processes and antiviral and interferon processes were enriched in most vaccines and vaccine types (**Fig. 5E)**. VV vaccines against COVID-19 and tuberculosis (MVA85A) were significantly (qvalue < 0.10) enriched with genes related to eosinophils, monocytes, and neutrophils (**Fig. 5E**).

## Discussion

This COVID-19 vaccination atlas comprehensively analyzes the dynamic and diverse immune responses generated by different vaccination strategies against COVID-19. These strategies include homologous and heterologous vaccination regimens, diverse vaccine types, and consideration of prior vaccination status at the time of infection, alongside the time elapsed since infection (**Fig. 1A**).

The analysis reveals that immune-related genes constitute a smaller proportion of the total DEGs than other biological processes, including cellular, metabolic, and regulation of biological processes (**Fig. 2A, Fig. S4)**. A limitation of this analysis by gene ontology of biological process is that some genes that are not annotated as immune play ubiquitous roles in cellular and metabolic processes present in immune cells. This can also be attributed to the inoculation of immunogens through syringe injection, which leads to local and limited tissue damage at the injection site. Indeed, intramuscular injections are prone to human errors, and questions remain regarding which injection site and procedure are optimal and safest ^31^. This vaccine administration issue and the molecular characteristics of different vaccine platforms might explain reactogenic behavior in some individuals. In the context of infection, this pattern illustrates the regulation of numerous genes influenced by viral infection cycle processes and the host response ^32,33^. This variability is primarily attributed to diverse response mechanisms among individuals, impacting disease progression or immunity^34^. However, this aspect will not be discussed in more details since it diverges from the primary objective of our study, which focuses on describing the transcriptome of the immune system. Nonetheless, it is noteworthy as it raises questions regarding non-immunological factors and their role in vaccination and disease development.

Moreover, it was found that these DEGs are highly shared between the studied conditions, encompassing both infection and vaccination conditions. In addition, a higher representation of immune-related genes was observed in the vaccination group compared to infection (**Fig. 2A**). Specifically, genes associated with inflammation, the complement system, neutrophil function, antiviral responses, and interferon pathways exhibit prolonged activation during infection, while adaptive immune-related genes are significantly downregulated (**Fig. S4-7**). In contrast, vaccination triggers the activation of chemokines and adaptive immune-related genes.

The surge in up– and downregulated DEGs in the early days after the second and third doses of BNT or MO indicates the ability to activate the innate immune response. Considering the rapid mRNA expression, post-inoculation might also trigger antigen-presenting cells to initiate an adaptive immune response ^35^. Notably, despite having the same mRNA platform as BNT and encoding the same spike amino acid sequence, the MO vaccine differs in nucleotide sequence due to codon optimization and the absence of pseudouridine modification^36,37^. However, whether this substantial increase is attributable to an immune response to the third dose, the technology, or other factors, such as LNP composition, remains unclear.

The heterologous vaccination regimen was also explored with different types of vaccines, namely ChAd and BNT, and BBIBP and ZF2001. It was found that it can contribute to the diversification of immune responses, address some of the technological limitations of the vaccine platforms, and consequently enhance protection against COVID-19.

The ChAd vaccine contains a chimpanzee adenovirus viral vector that shares epitopes with prevalent human adenoviruses, which can trigger an adaptive immune response against the vector^38^. It is therefore possible that the administration of multiple doses of the vaccine may generates antibodies targeting the vector, potentially limiting the delivery of genetic material, unlike mRNA vaccines ^39^. Hence, heterologous vaccination is a clinical and immunologically important option, allowing individuals who receive the complete homologous ChAd regimen to obtain better protection through vaccination with vaccines that use different technologies, particularly those based on mRNA ^16,40^. Nevertheless, the discussion regarding the continued efficacy of the ChAd viral vector in these individuals compared to other antigen vaccine formulations is crucial.

Heterologous mRNA vaccination regimens, especially those following the ChAd vaccine, demonstrate similarity to the second and third mRNA homologous and heterologous vaccinations. This can be advantageous for a vaccination regimen that is initiated by a VV vaccine and boosted by mRNA vaccines, potentially mitigating the impact of antibodies directed against the viral vector. However, the use of BNT as a third dose after two ChAd doses is mainly associated with antiviral and interferon responses as well as neutrophils. Additionally, as the samples were limited to the first day after vaccination in this condition, we could not assess the impact of these responses on the adaptive immune system.

The Covilo®/BBIBP vaccine is an inactivated virus vaccine formulated with alum adjuvant and evaluated in clinical trials with a homologous two-dose regimen(Wang et al., 2020; Al Kaabi et al., 2021; Zhang et al., 2022). The regimen of three homologous doses of BBIBP, specifically when given on days 7, 14, and 28, seems to present a limited immunological profile owing to the nature of the immune response to inactivated viruses^43^. In a study assessing the effectiveness of two doses of an inactivated influenza vaccine, employing a similar strategy to the development of the BBIBP vaccine, significant changes in gene expression were not observed one week after the second dose. Additionally, no considerable antibody titers were recorded, corroborating the BBIBP vaccine data^44^. However, in another BBIBP vaccine study, this limited immunological profile was not observed, in which a second boost induced more robust humoral and cellular immune responses detected after two weeks^45^. Despite the limited data regarding the efficacy of a third dose, it is evident that employing a homologous vaccination strategy with three doses of the inactivated BBIBP vaccine may offer limited immunological benefits. Nevertheless, the third dose demonstrated significant efficacy against the disease ^15^. It is important to emphasize the need to evaluate the immune transcriptome after the first two doses in samples from infected patients who have received two or three vaccinations^42^. Furthermore, investigating the early stages of vaccination, regardless of the number of doses administered, could provide valuable insights into identifying the genes linked to the innate immune response.

The Zifivax®/ZF2001 vaccine was not initially evaluated at the onset of the pandemic when the population had not yet been vaccinated. Instead, it was assessed as a booster dose in individuals who had previously received two doses of the BBIBP vaccine ^15^. As expected, in a heterologous regimen, vaccination resulted in a distinct gene expression profile when compared to the homologous vaccine ^11^. These findings suggest that immunological memory plays a pivotal role in the first-week post-vaccination, and antibodies generated by the inactivated vaccine recognize, opsonize, and trigger a response to the ZF2001 vaccine protein through the classical pathway of the complement system. Additionally, the original study revealed a significantly higher antibody titer than the homologous regimen, supporting the importance of heterologous vaccination ^15^.

Importantly, the findings demonstrate the significant contribution of complete vaccination regimens compared to natural infection and incomplete vaccination. Considering that infected patients with incomplete vaccination were hospitalized 10-11 days after vaccination with a single dose of BNT, infection must have occurred some days before hospitalization, and we anticipated that vaccination would not have protected these patients. In the PCA, individuals with prior-vaccination overlapped between the infected cluster and the vaccination cluster C2, which encompassed homologous vaccinations for both convalescent and non-convalescent individuals, along with healthy-vaccinated individuals (**Fig. 3A**). This overlapping was notable among individuals who were infected after a complete vaccination regimen with BNT. Conversely, an incomplete vaccination regimen was associated with infection, suggesting a limited protection of vaccination administered right before infection. We found that in infected patients with no prior vaccination or with incomplete vaccination the genes related to innate immune cells and processes were highly activated and persisted for several days, especially IFN1 and IFN2, neutrophils, and APCs. Interestingly, bacterium-directed PRRs (TLR4 and NOD) were also highly expressed in these conditions, suggesting that these patients were also coinfected by bacteria, which is a risk factor for hospitalization^46^.

To provide a model of key genes that describe infection and the different types of COVID-19 vaccines, we trained a classification random forest model with 244 genes selected from the PCA. Our model identified the 10 genes that are most important for classification, with high accuracy (91.2%) and sensitivity (ROC AUC of 99.0%), and low OOB ratio (8.6%). The genes that classified infection were associated with transcriptional regulation (GATA3, ZNF3, KMT2A, and ASXL1), antiviral response (SP100), and granzymes (GZMM); vaccine types were marked by ARPP (ITGAM, ACTG1, LGALS3) and T-cell response (STAT5B) (**Fig. 4B**).

In addition, we compare COVID-19 vaccines with other vaccines targeting different pathogens, as outlined in the Vax MSigDB dataset (**Fig. 5C**). This comparison is crucial for understanding how new-generation vaccines, such as mRNA-based and viral-vectored vaccines, relate to classical and consolidated vaccine technologies. Given the pivotal role of the innate immune response in the early stages of both vaccination and infection, the overlapping genes identified likely contribute to the innate immune system ^47^. In particular, bias in FCER1G towards non-COVID-19 vaccines shows a predominance of Th2 response, presumably due to the widespread use of alum adjuvants, a known Th2 response activator that is used in the formulation of BBIBP and ZF2001 ^48^. This comparison was notable with influenza IN vaccines and LA vaccines against yellow fever and smallpox, which share common DEGs with COVID-19 vaccines in the initial weeks, especially those related to the innate immune system. Additionally, LA vaccines serve as a valuable reference for a robust immune response, mimicking the wild-type virus infection. The observed overlap of LA vaccines with COVID-19 vaccination conditions adds an intriguing dimension to our findings. The diversity in the immunome in vaccines sharing the same technology may be attributed to the specific and varied nature of the adaptive immune system and other technical limitations of the MSigDB Vax dataset. It primarily relies on transcriptomic data from microarrays, which predates the widespread adoption of high-throughput RNA sequencing. In addition, this dataset lacked gene expression fold-change values, highlighting a constraint in our analysis.

Although this study provides valuable insights, there are notable limitations regarding the datasets and transcriptomic analyses utilized. Specifically, the use of only bulk RNA-seq data restricts our understanding of the dynamic and heterogeneous behavior of individual immune cells. Furthermore, collecting vaccination and infection samples at different time points presents a limitation in describing the chronological dynamics of the immune response. Specifically, this difference restricts the characterization of the adaptive immune response in vaccination, predominantly assessed within a one-week period. There are additional challenges in analyzing third-dose vaccinations with BNT, MO, and ChAd, which were assessed on the first-day post vaccination, primarily constraining the analysis to the innate and complement immune systems. These restrictions remain significant despite their interconnectedness with the adaptive immune responses elicited by previous doses. In addition, the analysis of BBIBP and ZF2001 vaccines is restricted by the limited time points on the days during the first week, crucial for the innate immune response, and also did not allow us to compare with the preceding doses. Moreover, the observed gene expression pattern was in agreement with the immunological finding of a study, but was contradictory to another(Aydillo et al., 2022; Ying Chen et al., 2023). Exploring how different vaccine technologies contribute to protecting infected vaccinated individuals, as observed in BNT-vaccinated individuals, would provide further insight.

To improve analysis and interpretation by incorporating supplementary data sources, such as antibody titers, neutralizing antibodies, T cell counts, and clinical trial data, would greatly enhance our understanding. Although sex, age, and VOCs data were present, we did not adjust the contribution of these variables to the immunological signatures described. Adopting a multidimensional approach that integrates various datasets and immunological methodologies would offer a more comprehensive understanding of the immunological landscape after vaccination.

## 3. Methods

### Data curation and processing

The selected datasets were first curated using available metadata from Gene Expression Omnibus (GEO) and supplemented with information obtained from pertinent publications (**Fig. S1-3, Table 1)**. We included studies related to SARS-CoV-2 immunization that provided detailed and precise methodology, involved convalescent and non-convalescent patients with or without prior vaccination, and used bulk or single-cell RNA-seq PBMC-derived samples. We excluded studies that lacked methodological descriptions or references, especially concerning sample collection timing between doses, involved TCR and BCR repertoire sequencing, included patients with distinct medical conditions such as autoimmune diseases and cancer, or lacked standardized count matrices. The count matrices for each dataset were obtained from the Gene Expression Omnibus (GEO) ^49^ and their metadata from the R package GEOquery ^50^. To standardize the gene identifiers as HGNC symbols, we used the R package biomaRT ^51^. For reprocessing of reads in the GSE206023 study, FASTQ files were downloaded directly from GEO under the accession number GSE206023. **The details of the dataset and its accession numbers can be found in Supplementary Table ST1.** The average number of reads per sample was approximately 36 million after trimming, which can also be found in the supplementary material. Quality control of raw and trimmed reads was performed using FastQC v.0.11.8 (Andrews, 2010). Trimming of the adapter content and quality trimming was performed using Trimmomatics v0.36, with the following settings: LEADING 20 TRAILING 20 SLIDINGWINDOW 4:25 MINLEN 31, and the Kallisto index was built with reference transcriptome GRCh38 (Ensembl) with a k-mer length of 31 using the Kallisto v.46.0 programs, and the abundance of the transcripts was quantified using the Kallisto pseudo-alignment, which provides estimates of transcript levels ^17,52,53^. Subsequently, we used the tximport R package to summarize count estimates at the gene level ^54^.

### Differential gene expression analysis

Differential gene expression analysis was performed independently for each dataset using the R package DESeq2 ^55^. We used the Wald test with the interaction terms vaccine and time point. DEGs were filtered based on the Benjamini-Hochberg adjusted p-values (padj <0.05). We classified up and downregulated genes by log2 fold change (–1 < L2FC > 1, respectively).

### Gene set enrichment analysis

The ontology of biological processes within the immune system [“immune system process” (GO:0002376)] was obtained from the GO.db package ^56^. To facilitate subsequent analyses, biological processes associated with the immune system were categorized into three distinct groups: “Immune system,” “Immune subsystem,” and, when related to specific cells, classified under the “Immune cell” category. The VAX MSigDB database was obtained via the MSigDB portal and manually annotated with the available information on the dataset. We used only peripheral blood mononuclear cell-derived samples. The enrichment analysis of DESeq2 results was conducted with ImmuneGO using the Enricher and Clusterprofiler packages for over-representation analysis (ORA) and gene set enrichment analysis (GSEA) ^57^. Heatmaps were visualized using the ComplexHeatmap package in R and Morpheus (https://software.broadinstitute.org/morpheus) ^58,59^.

### Principal component analysis and machine learning classification

PCA was performed using L2FC values of immune-related DEGs, through the factoextra R package ^60^. For cluster analysis, we used the k-nearest neighbors (KNN) algorithm. To optimize the efficiency of our random forest classification, we initially filtered genes based on their cosine squared values in the first two principal components, setting a threshold above 0.5. We then constructed our random forest classifier by leveraging the tidymodels package (Kuhn & Wickam, 2020). Our dataset was partitioned into training (70%) and testing (30%) sets, with stratification for the “type” variable and upsampling to address class imbalance. Employing cross-validation, we fine-tuned the model’s hyperparameters using a grid search method within the tidymodels framework. Performance evaluation metrics, including AUC, ROC, OOB, and accuracy, guided the optimization of hyperparameters, including mtry and min_n. Finally, the optimized random forest model was trained on the entire training dataset and subsequently assessed on the testing dataset to evaluate its predictive performance.

## Data Availability

All data produced are available online at

https://github.com/wapsyed/covidvax_atlas

## Data and code availability

Raw and processed data, as well as codes used are available on the project Github repository (https://github.com/wapsyed/covidvax_atlas).

## Author Contributions

WAPS, DLMF — Conceptualization and manuscript writing;

LV, AL, NC, EC, JSD — Manuscript contributions;

WAPS, DLMF, SZP — Data analysis;

OC, HIN, LFS, ISF, GW, RDC, NOSC, HDD, HDO, ECS, JEK – Review and editing;

GCM, OCM, HIN – Supervision, editing, and approval of the final version.

## Funding

We thank the São Paulo Research Foundation (FAPESP grants 2019/14526-0 and 2020/05146-7 to GCM; 2018/18886-9 to OCM; 2023/07806-2 to ISF; 2020/16246-2 and 2023/13356-0 to DLMF; 2019/01255-9 and 2021/03684-4 to RDC) for financial support. We acknowledge the National Council for Scientific and Technological Development (CNPq) Brazil (grants: 309482/2022-4 to OCM and 102430/2022-5 to LFS).

## Conflicts of Interest

The authors declare that they have no competing interests.

## Supplementary Material

### Supplementary Figures

**Fig. S1.**
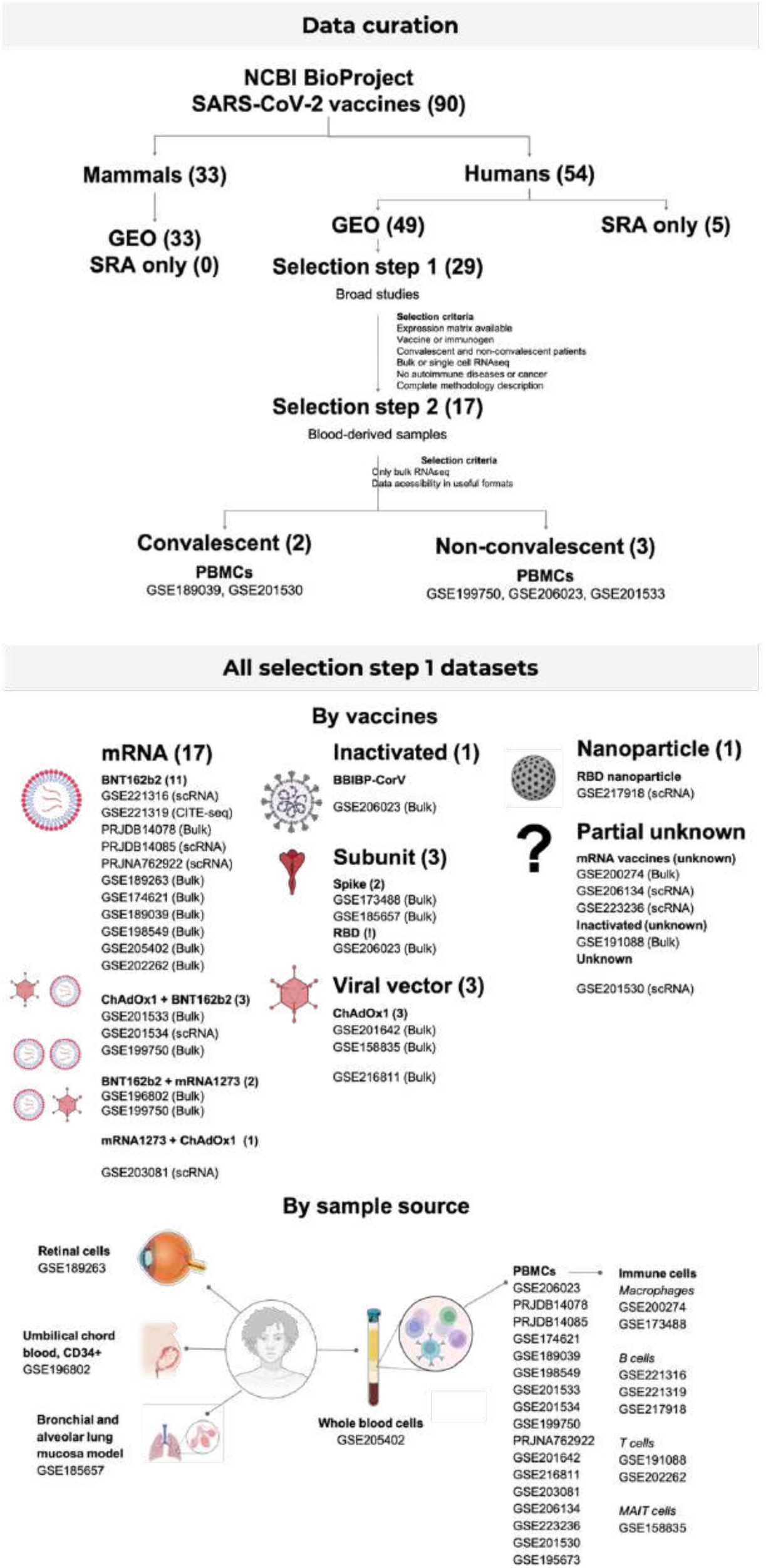
Data curation. Datasets were obtained from NCBI BioProject and curated. All datasets, including the selected and non-selected, were classified by vaccine technology and vaccination regimen, and by sample source.

**Fig. S2.**
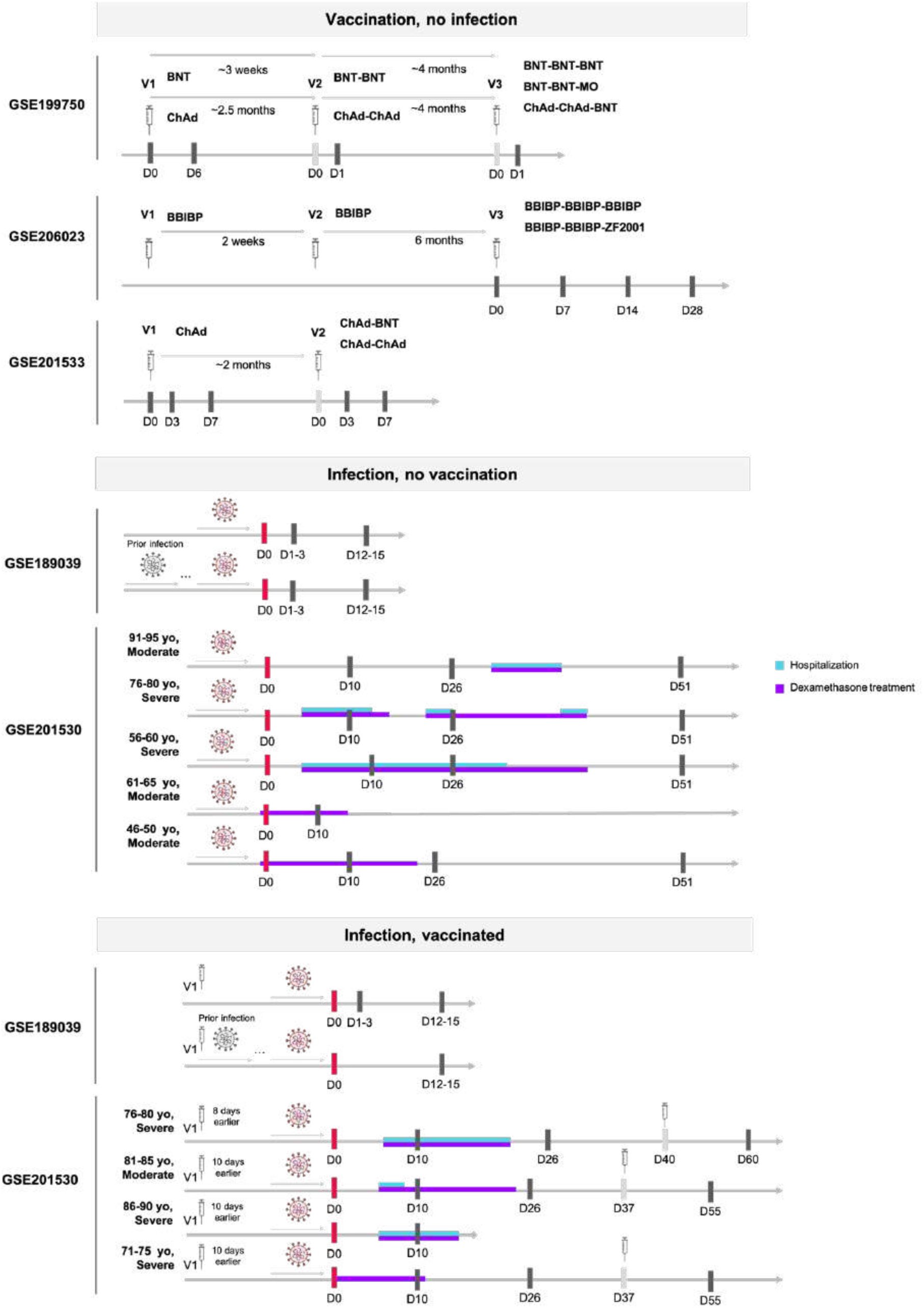
Description of the selected datasets. The datasets include healthy vaccinees without prior infection, infected patients without vaccination, and patients with prior full or incomplete vaccination. Sample collection timelines are shown as black lines, with red lines indicating the first sample collection during hospitalization and gray lines representing events without sample collection. Days post-infection or vaccination are depicted, with blue lines representing hospitalization periods and purple lines indicating dexamethasone treatment.

**Fig. S3.**
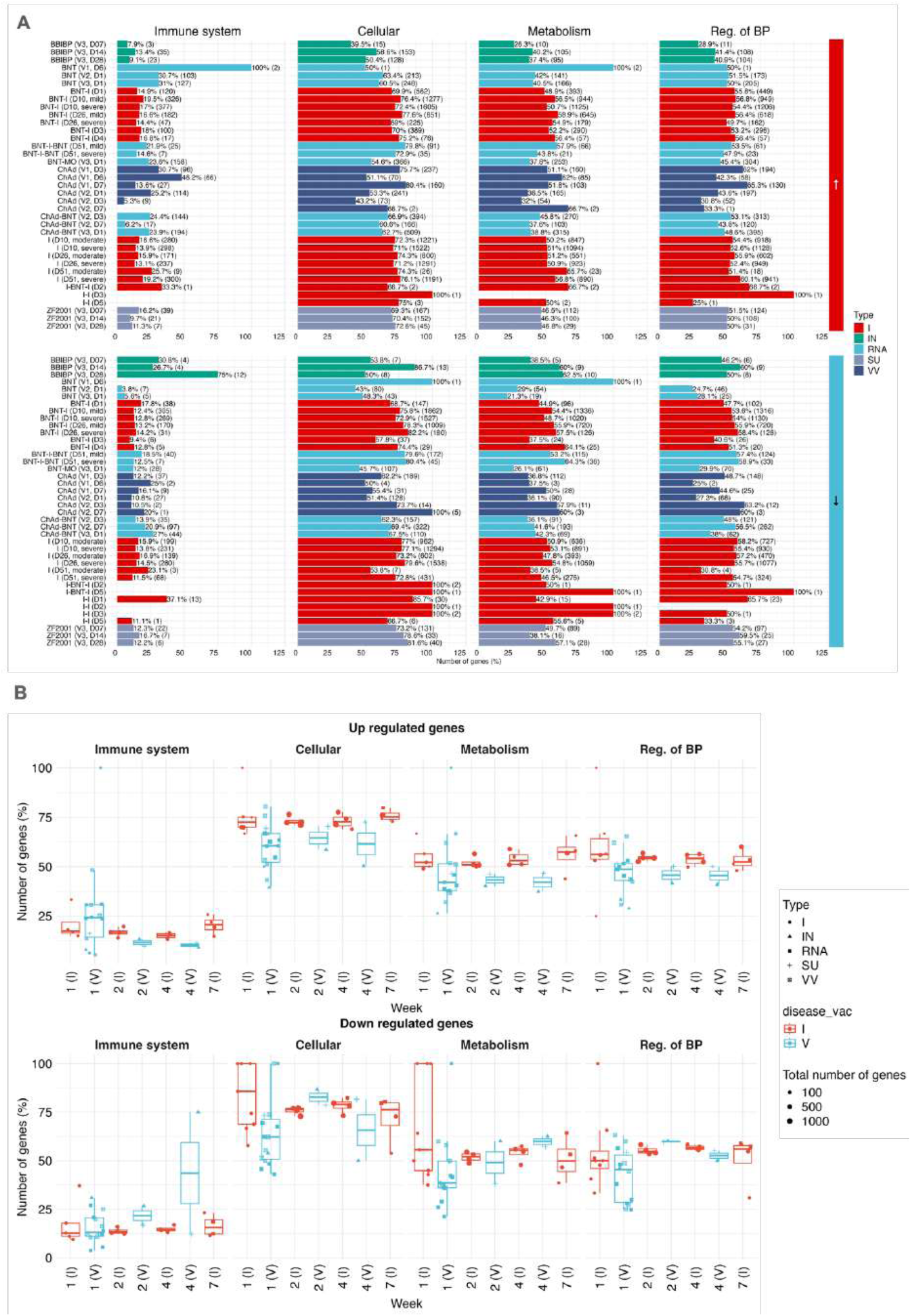
DEGs were categorized as up-(A) and down-regulated (B) with the absolute and relative number of genes, and grouped based on infection (red) and vaccination (blue) conditions. The conditions were then plotted with the relative number of genes corresponding to their roles in essential biological processes, namely immune system, cellular processes, metabolism, and regulation of biological processes, respectively. Shapes correspond to type, colors to disease or infection, and size to total number of DEGs. I: Infection; IN: Inactivated; RNA: mRNA vaccines; SU: Subunit; VV: Viral vector.

**Fig. S4.**
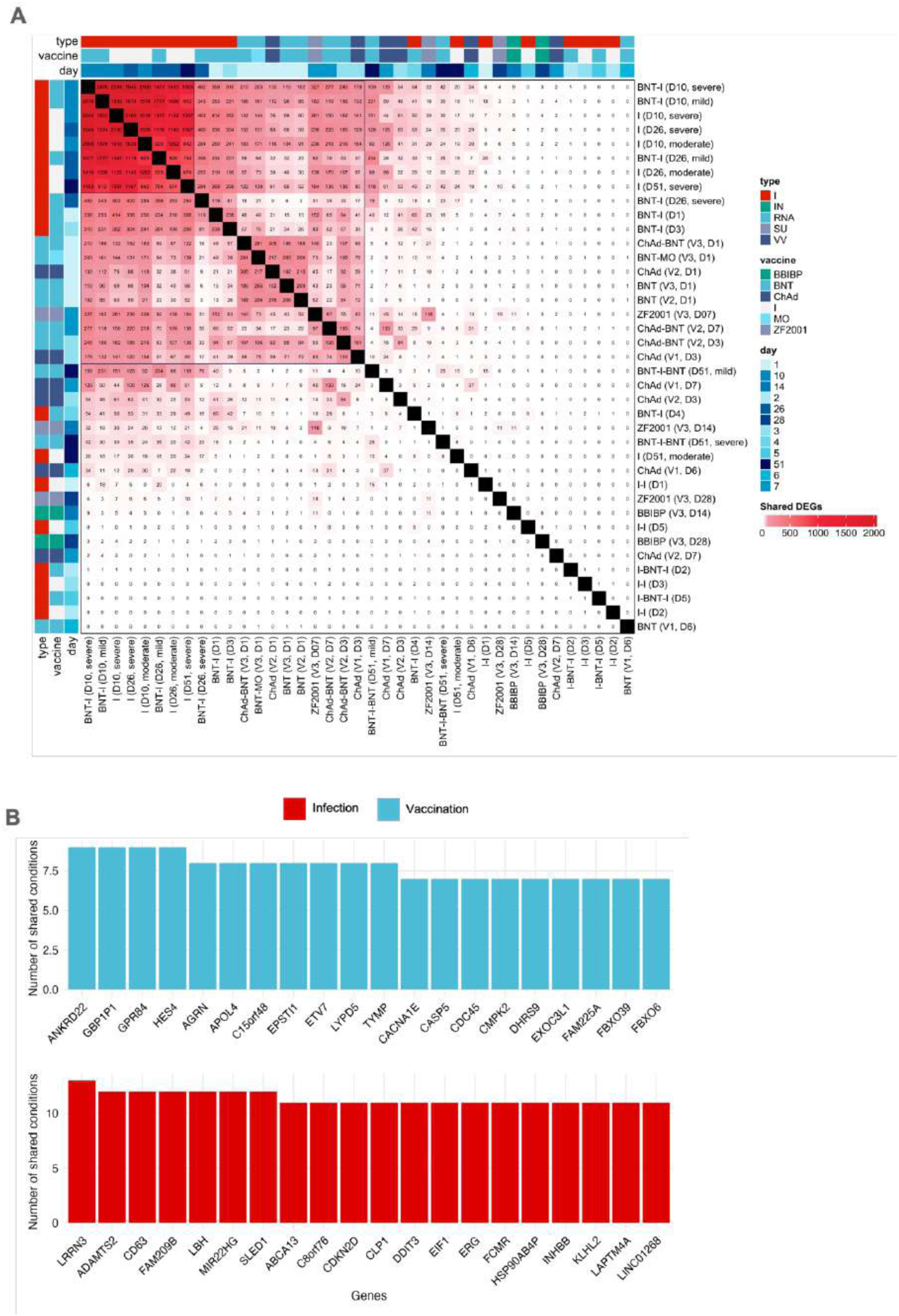
A. Shared non-immune-related DEGs between infection and vaccination. B. Twenty most shared immune-related genes. BBIBP: BBIBP-CORV; BNT: BNT162b2, ChAd: ChAdOx-1; MO: mRNA-1273; I: Infected; V: Vaccinated; IN: Inactivated; RNA: mRNA vaccines; SU: Subunit; VV: Viral vector.

**Fig. S5.**
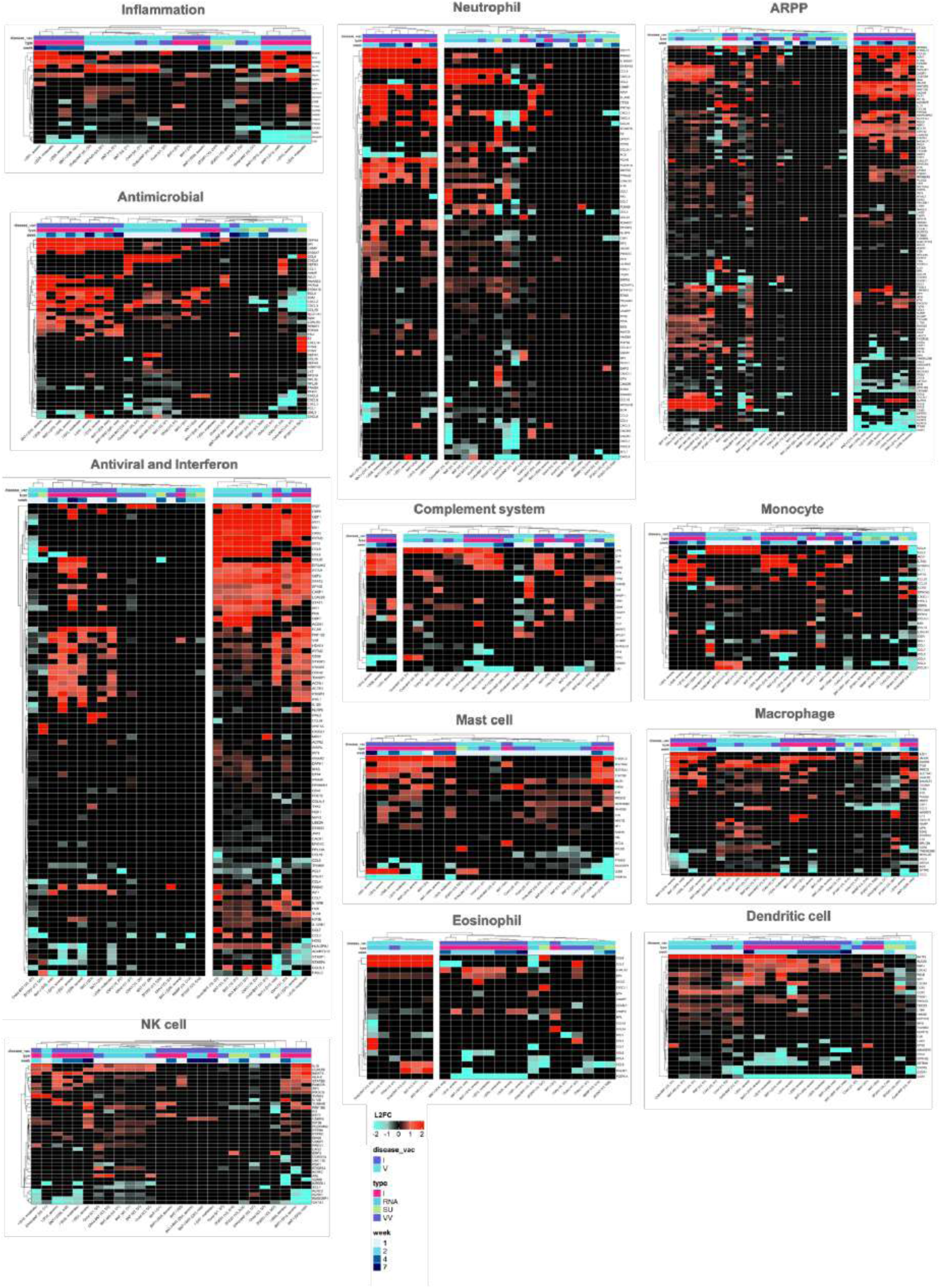
Genes related to complement system, antimicrobial response, and innate immune system processes and cells. I: Infection; IN: Inactivated; RNA: mRNA vaccines; SU: Subunit; VV: Viral vector.

**Fig. S6.**
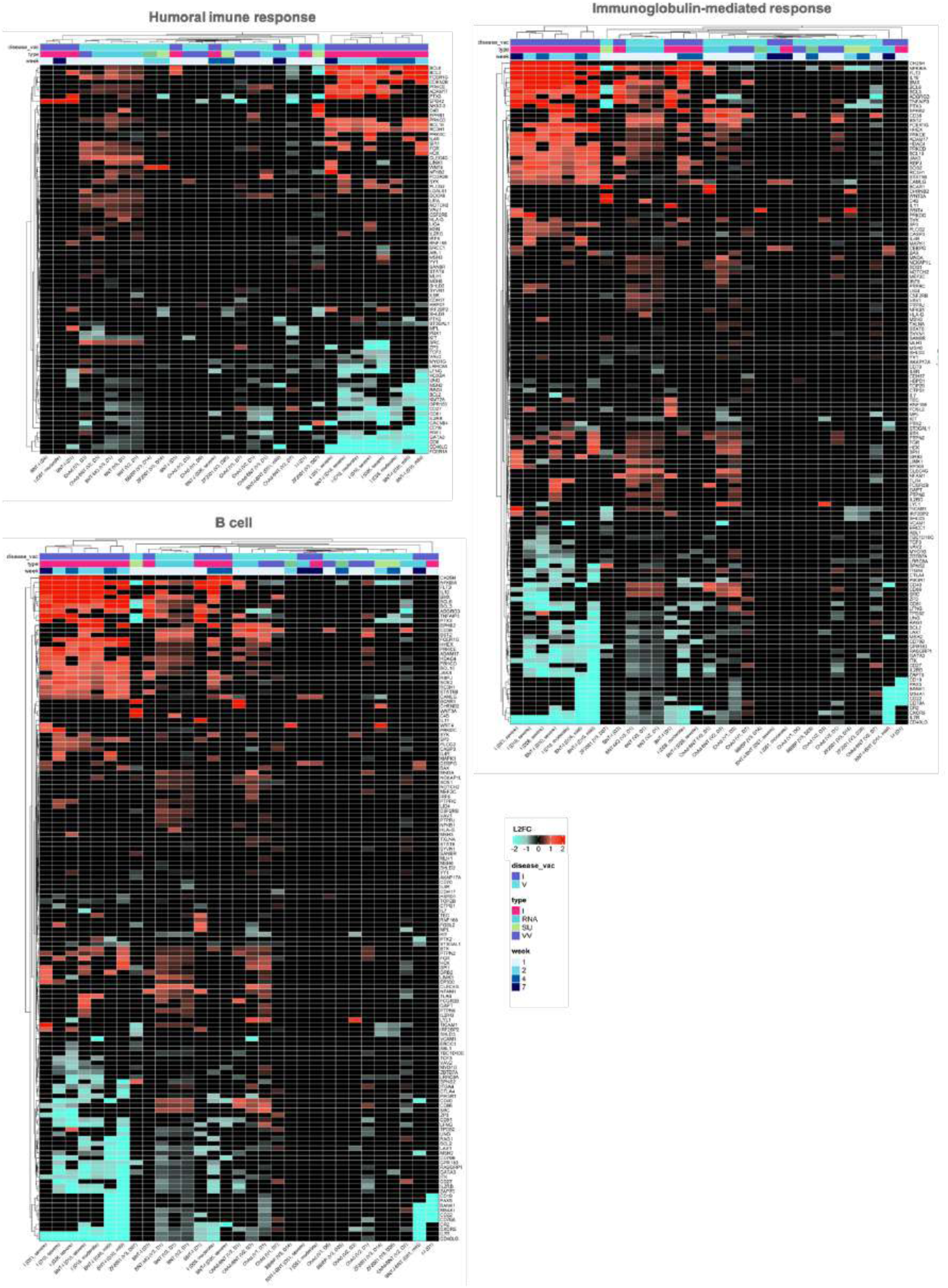
Genes related to humoral adaptive immune system, including B cell processes and immunoglobulin-mediated response. I: Infection; IN: Inactivated; RNA: mRNA vaccines; SU: Subunit; VV: Viral vector.

**Fig. S7.**
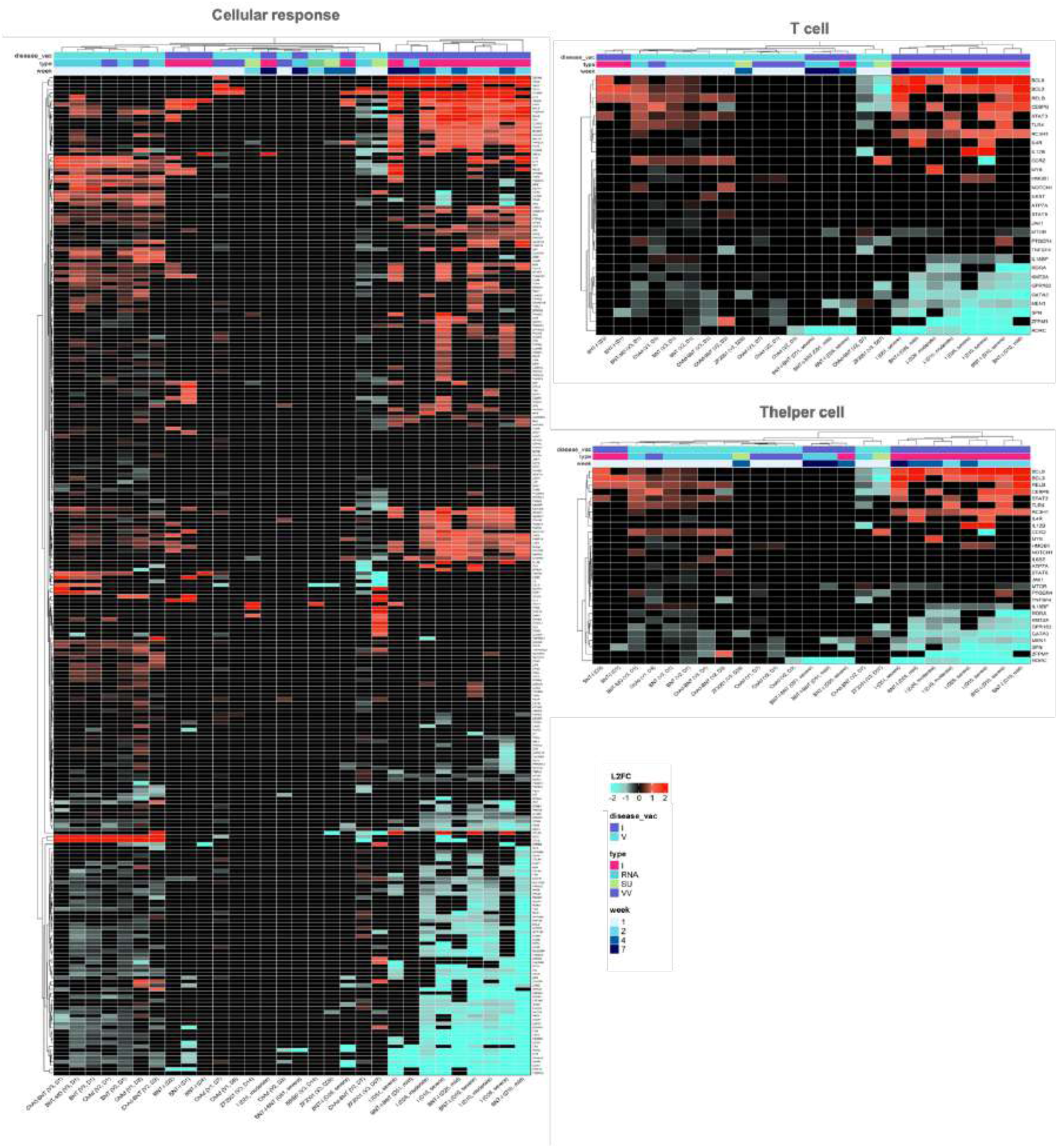
Genes related to the cellular adaptive immune system, including cellular response and T– and T helper cell processes. I: Infection; IN: Inactivated; RNA: mRNA vaccines; SU: Subunit; VV: Viral vector.

**Fig. S8.**
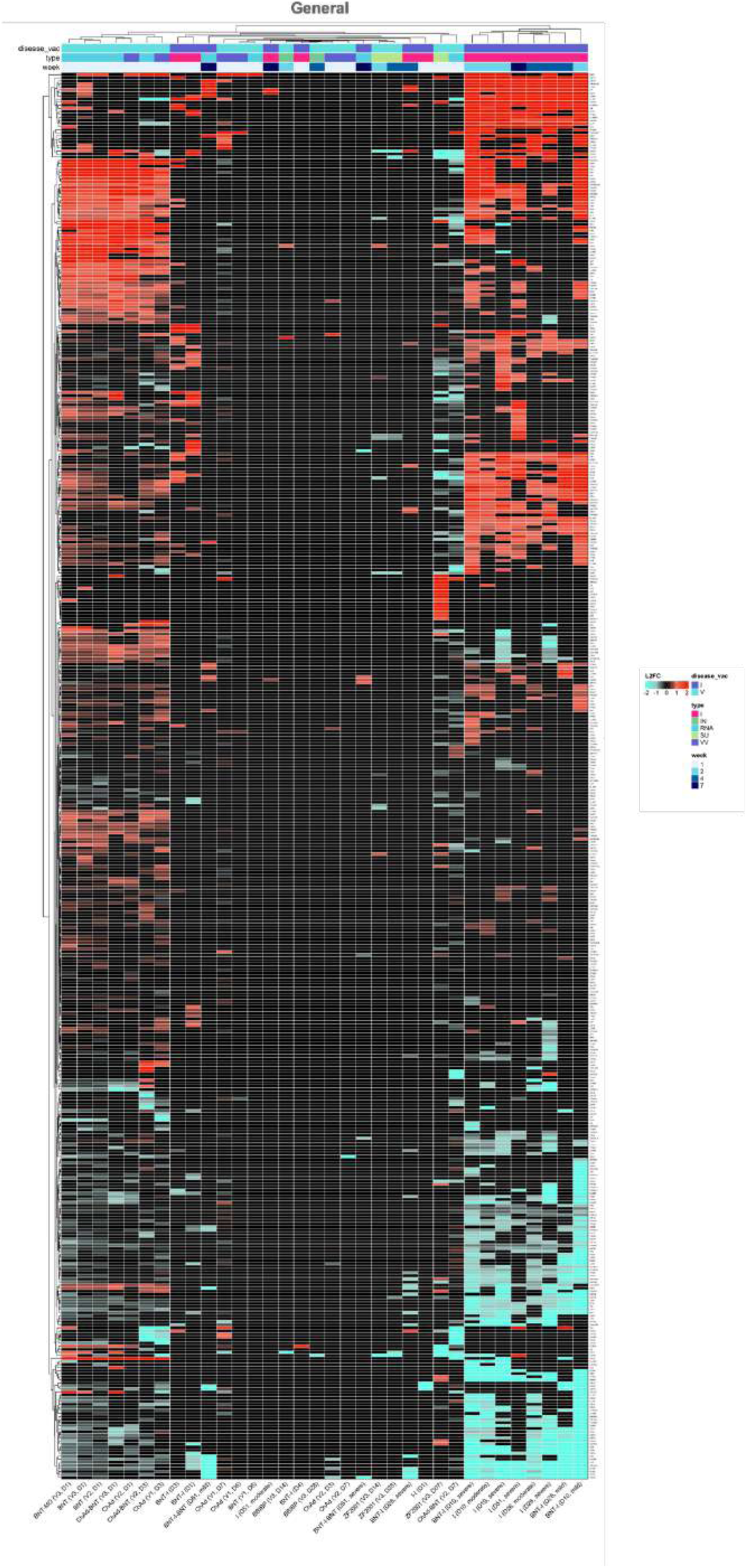
General leukocyte processes. I: Infection; IN: Inactivated; RNA: mRNA vaccines; SU: Subunit; VV: Viral vector.

**Fig. S9.**
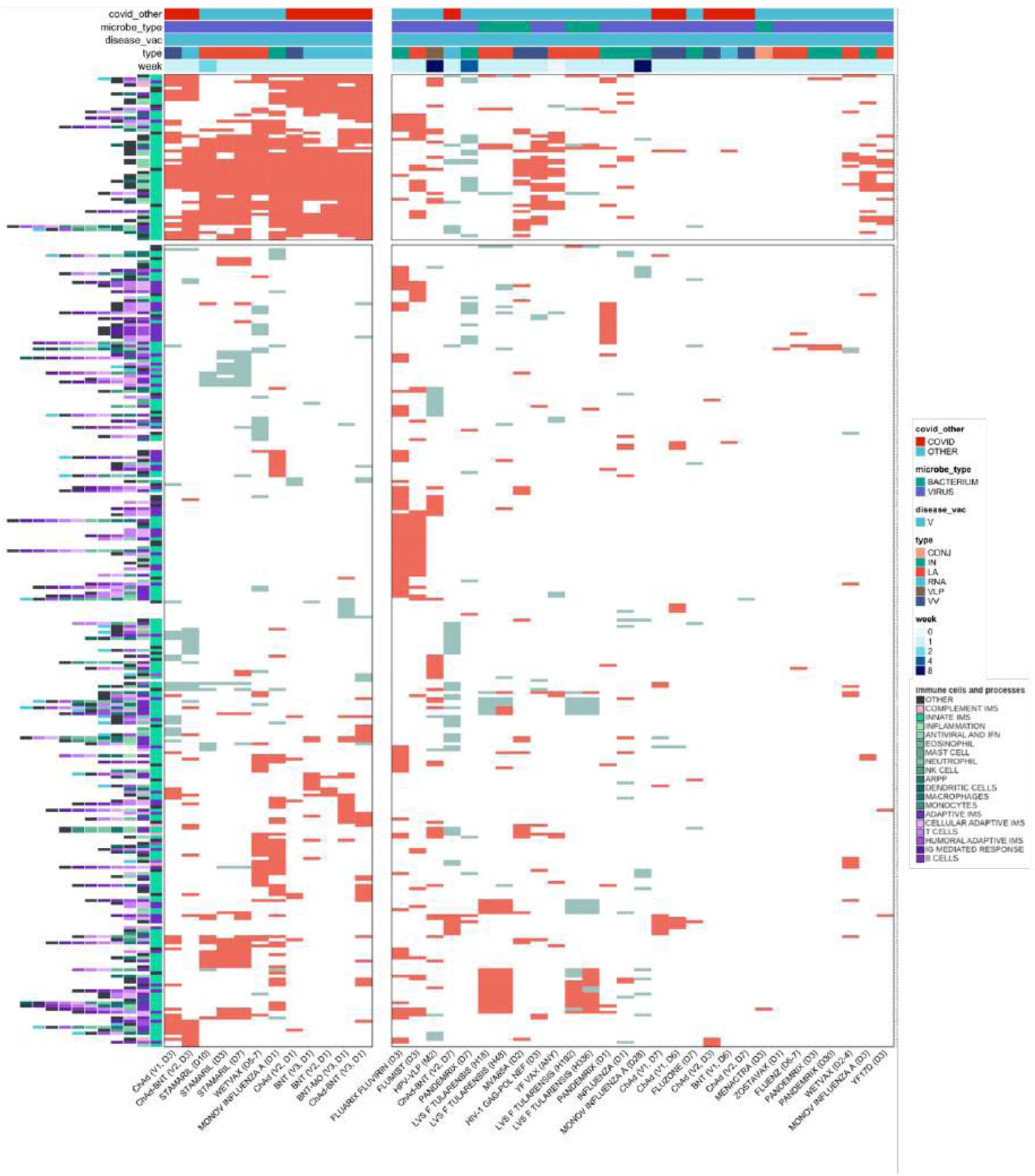
Gene expression in vaccination against COVID-19 and other pathogens. I: Infection; IN: Inactivated; RNA: mRNA vaccines; SU: Subunit; VV: Viral vector; CONJ: Conjugated; LA: Live-attenuated; VLP: Virus-like particle.

## Notes

### Competing Interest Statement

The authors have declared no competing interest.

### Funding Statement

We thank the Sao Paulo Research Foundation (FAPESP grants 2019/14526-0 and 2020/05146-7 to GCM; 2018/18886-9 to OCM; 2023/07806-2 to ISF; 2020/16246-2 and 2023/13356-0 to DLMF; 2019/01255-9 and 2021/03684-4 to RDC) for financial support. We acknowledge the National Council for Scientific and Technological Development (CNPq) Brazil (grants: 309482/2022-4 to OCM and 102430/2022-5 to LFS).

### Author Declarations

Gene Expression Omnibus (GEO) identification: GSE199750 GSE201530 GSE189039 GSE201533 GSE206023

### Summary of Updates

The author list, the figures and the manuscript were updated.

